# Quantifying the impact of social activities on SARS-CoV-2 transmission using Google mobility reports

**DOI:** 10.1101/2024.01.03.24300755

**Authors:** Felix Günther, Hilde Kjelgaard Brustad, Arnoldo Frigessi, Tom Britton

## Abstract

We developed a state-space model to investigate which social behaviours had biggest impact on the spread of SARS-CoV-2. The analyses were based on reported hospitalizations, together with information on vaccinations, weather data, virus strains and, most importantly, Google mobility reports on 4 different types of social activities. While our new approach is general, we studied Sweden and Norway on a regional level over 75 weeks, and the major regions of Berlin and Bavaria in Germany over 10 months. Most results are shared for all three countries: Activity in four social settings explain between 40-60% of all infections; Public transport appears as an important setting for infections in all countries; and the transmission potential drops by 40-50% during the summer as compared to the winter peak. However, the analyses for Germany differ in that *Retail and recreation* is the other setting dominating transmission whereas it is contacts at the *Workplace* in Norway and Sweden, showing how our model is able to adapt to specific cases. Transmissions not captured by the Google data may happen in other settings, in particular in households. The statistical model has a deterministic time and region specific transmission rate with an additive component for the four Google settings, and a multiplicative part taking seasonality and circulating virus strains into account. Inference is performed in a Bayesian setting using Stan.

## 1 Introduction

How individuals move and interact in the society affects the transmissibility of an infectious disease. In mathematical modeling of infectious diseases, this is commonly reflected by defining the transmissibility of an infectious disease as proportional to the product of the number of contacts between infectious and susceptible persons and the probability of infection per contact at a certain point in time. Depending on the pathogen, different modes of transmission exist. For respiratory viral diseases, like SARS-CoV-2, four central modes of infection are often considered: direct (physical) contact, indirect contact (via fomites), (large) droplets and (fine) aerosols (14). Importance of these modes varies between pathogens and is often not entirely certain, remaining a point of scientific debate. From a theoretical perspective this implies that a somewhat abstract quantity is relevant to describe the spread of infectious diseases systematically: an effective number of contacts that can lead to infection and includes all possible transmission routes.

Information on the (temporally and geographically varying) number or intensity of such contacts is of great importance for public health autyhorities and disease control. On the one hand, they can help to assess the potential for spread of a pathogen and thus the dangerousness of the situation; on the other hand, they allow to assess the effectiveness of any measures taken. However, in practice, the (average) number of contacts relevant for transmission is unfortunately unknown and notoriously difficult to measure.

During the SARS-CoV-2 pandemic, different and partly novel data sources were used as proxies to measure (changes in) contact frequencies or, more generally, social activity within populations: contact surveys, mobility data collected, for example, by mobile phone providers, and other indicators resulting from repurposing routinely collected location data by various, often private actors. Such data sources have been used in various studies in the context of the SARS-CoV-2 pandemic focusing on different questions. For example, the Norwegian Institute of Public Health (FHI) has embedded Telenor mobile phone data in its meta-population models as a proxy for mobility between regions (1). Badr et al. (5) used aggregated mobile phone data to create a social distance metric for each US county and investigated its correlation with the growth-rate of Covid-19 case counts. Mobile phone data have also been used to create detailed networks of how individuals in a population move between different points of interest and how they interact in different parts of the society (4; 7). Fritz et al. (12) used data from Facebook’s Data for Good program to investigate how regional differences in mobility patterns and friendship proximity affected the spread of COVID-19 in Germany. It is, however, oftentimes not entirely clear how well these data sources represent changes in relevant contact frequencies, which is particularly important when being interested in directly using these indicators, and their variation in time and space, for surveillance.

An important and publicly available data source on population behaviour during the SARS-CoV-2 pandemic were the Google community mobility reports (GCMR) (2). This quantity was defined as the percentage increase or decrease in activity levels relative to a pre-pandemic baseline level in six different settings: grocery and pharmacy, retail and recreation, transit stations, parks, workplaces, and residential areas. The activity data are generated from mobile devices with a Google account where the location history setting is enabled. The GCMR data were collected and updated until 2022-10-15 and are freely accessible from Google in a fine-grained resolution in time and space. More details are given in the Materials and Methods section. These data were used in numerous studies investigating different aspects of SARS-CoV-2 spread. Cot et. al (8) combined the GCMR and Apple Maps Mobility Trends Reports (AMMTR) to quantify the impact of social distance on infection rates in European countries and in the US and to identify a time scale between the start of social distancing and the reduction in infection rates. Their results indicate a reduction of 20%-40% for European countries and 30%-70% in the US, during the second wave of the Covid-19 pandemic. A study by Nouvellet et al. (15), which combined a renewal process with a parametrization of the reproduction number as a function of GCMR and AMMTR, indicated a change in association between human behaviour and transmissibility over time. Basellini et al. (6) reduced the GCMR to a single mobility index by time and region in England and Wales through multilinear principal component analysis and linked estimates of excess mortality to this mobility index. A study by Yilmazkuday (23) used a difference-in-difference design with the different settings from the GCMR as a continuous treatment, to investigate its impact on change in COVID-19 cases and deaths for 130 countries. The results suggested that a 1% weekly increase in the GCMR residential setting led to about 70 less cases on average across all countries in the study. A 1% decrease in the work setting, transit setting or retail and recreation setting was estimated to give 18, 33 and 25 less cases, respectively.

The aim of our work is to investigate to what extent the GCMR data are suitable to explain changes in transmission dynamics during the SARS-CoV-2 pandemic and whether the different activity categories of the GCMRs can be compared to make statements about which areas of social activity were particularly relevant for disease spread.

To achieve this, we have created a meta-population model, which we used to describe the disease spread in individual regions, and linked this model to a principled model of the association of GCMRs with contact frequencies among individuals in specific settings. Importantly, we carefully modeled the GCMR indicators, which refer to a pre-epidemic reference period and therefore need a accurate mathematical treatment, to make interpretation possible. In our main analysis, we applied the model at a relatively fine spatial resolution for Norwegian and Swedish regions in the period from February 2020 to July 2021. The model allowed us to take into account changes in the probability of infection due to seasonality and new virus variants, as well as the start of the vaccination campaigns in 2021. Our proposed model was flexible and adaptable, which we illustrate in a second analysis based on publicly available data in two different regions of Germany.

## 2 Results

### 2.1 Modeling Google Community Mobility Reports and transmission dynamics of SARS-CoV-2

A central part of our work has been to develop a theoretically well-founded model in which the measurements of the GCMR can be brought into a interpretable relationship with the transmission dynamics of SARS-CoV-2. In order to take into account general aspects that regulate the spread of an infectious disease - for example, the time-varying proportion of susceptible individuals in the population and the age dependent vaccination against SARS-CoV-2 in our study period - we work with an extended SIR model per region with discrete daily time-steps. In such models, the transmissibility of the virus in a given region *r* on day *t* is determined by a parameter *β*_*t,r*_, which represents the average number of secondary infections per infected individual in region *r* on a given day *t*, in a hypothetical fully susceptible population. Based on theoretical considerations, this parameter can be further factorized into two parts: the average number of contacts each infected individual has and the probability of infection per contact.

An important assumption of our model is that the level of activity in a GCMR setting is linearly related to the number of contacts in that setting, relating the GCMR 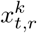 to the number of contacts 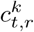, in setting *k* as

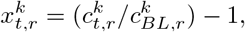

where 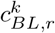 denotes the unknown average number of contacts an infectious individual have in setting *k* at a pre-pandemic baseline time. The quantity 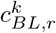 is included because the GCMR only provide information on the activity level in a setting *k* and region *r* compared to the region and setting specific pre-pandemic baseline. Furthermore, we assume that the contacts in the different settings add up to the total number of contacts. The contribution of a GCMR setting *k* to the daily transmissibility parameter *β*_*t,r*_ is then defined as

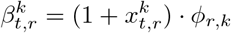

where *ϕ*_*r,k*_ is the product between the region and setting specific baseline number of contacts 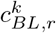 and the probability of infection upon contact 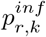. Note, that this relation between activity level and the number of contacts as well as the infection probability per contact can differ from setting to setting. Furthermore, additional factors can change the probability of infection per contact, e.g., seasonal factors or changes in infectiousness due to mutations towards new variants. Therefore we propose to model the transmissibility parameter *β*_*t,r*_ as a function of the GCMR and other multiplicative factors in the following way:

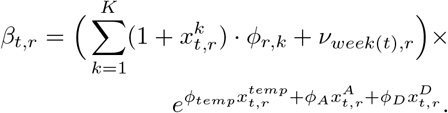

Here, 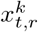 are the measurements of the seven day average of the *k*-th GCMR on day *t* in region *r* scaled such that 0 corresponds to the baseline level, 1 corresponds to a doubling in the number of visitors and, e.g., −0.5 corresponds to a reduction in activity by 50%. Further data are 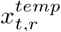, the temperature difference on day *t* compared to February 21, 2020, in degrees Celsius, as well as 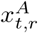 and 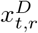, the share of the Alpha and Delta Variant of Concern (VoC) among all infected at day *t* in region *r*.

We estimate the parameters *ϕ* and *ν* by fitting the model to weekly hospitalization counts in our observation period: the parameters *ϕ*_*r,k*_ capture the relevance of the *k*-th mobility report for SARS-CoV-2 transmission. At the unknown pre-pandemic activity level in mobility setting *k* we expect in region *r* on average *ϕ*_*r,k*_ secondary infections per infectious individual per day due to population activity and corresponding contacts captured by the corresponding GCMR mobility report.

If the GCMR activity level in a setting is reduced, e.g., by 50%, the model assumes a linear reduction of the infections contracted in this setting by 50% as well. This linearity assumption appears plausible for the four settings Retail and recreation, Grocery and pharmacy, Transit stations and Workplaces. However, for the Residential setting, a linear relation between time spent at home and the average number of secondary infections appears implausible. Within a household contacts are so intense and persistent, and the number of household members limited, that more time spent at home over a ceetain threshold has no significant impact on infections. Additionally, there is a strong negative correlation between time spent in the Residential setting and the others, making fitting harder. For these reasons the Residential setting was left out of the analysis, and so was the Park setting because very little transmission happened outdoor. Therefore, we restrict our model to the four GCMR mentioned above.

Since the four GCMR settings do not contain all infection sites and are most likely insufficient to capture all relevant changes in contact frequencies over time, we include an additional region- and week-specific parameter *ν*_*week*(*t*),*r*_ that accounts for secondary infections outside the four settings considered.

The effects of the temperature difference, 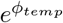, and VoC’s, 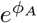 and 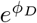, correspond to multiplicative increase of the transmission probability per contact. We use the temperature as a proxy for changes related to seasonality. The effects of the VoC’s quantify an increase in transmissibility of those virus variants per contact compared to the wildtype. We assume that these multiplicative effects are the same in each considered region. A more detailed motivation and description of the model, as well as transformations of the estimated parameters that we use for summarising our results are given in the Materials and Methods section.

### 2.2 Model fit to hospitalization data

For Norway and Sweden, we fitted our model to weekly hospitalization counts of Swedish and Norwegian regions accounting for age-prioritised vaccination roll-out in the beginning of 2021 and the emergence of the Alpha and Delta VoC’s. Estimation was performed in a Bayesian setting using Hamiltonian Monte Carlo in a custom implementation of the model in Stan (19) using the CmdStanR interface (13). Detailed information on the definition of the region-level extended SIR model, the likelihood for the weekly hospitalization counts derived from the SIR model, as well as the fixed and estimated parameters and their priors used during estimation can be found in the Materials and Methods section. We fitted separate models, one for joint modeling of the 11 Norwegian regions (*fylke*) and one for the 21 regions (*län)* of Sweden.

Visual investigation of trace plots indicated convergence of the four independent MCMC chains per model to a joint posterior distributions, furthermore the standard convergence diagnostic 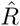 provided by Stan (20) was smaller than 1.01 for all sampled parameters. Fig. 1 shows that our highly flexible model is able to capture the development of weekly hospitalization counts well, visualised by Oslo in Norway and Stockholm in Sweden. Panel A illustrates the model fit, Panel B shows the estimated effective reproduction number and Panel C the contribution of the different GMR categories over time. The flexibility of the model stems from the large amount of parameters that are estimated, in particular the region specific weekly number of secondary infection not captured in the GCMR setting, *ν*_*week*(*t*),*r*_. Supplementary figures S19 - S24 provide similar plots for each Norwegian and Swedish region.

**Fig. 1:**
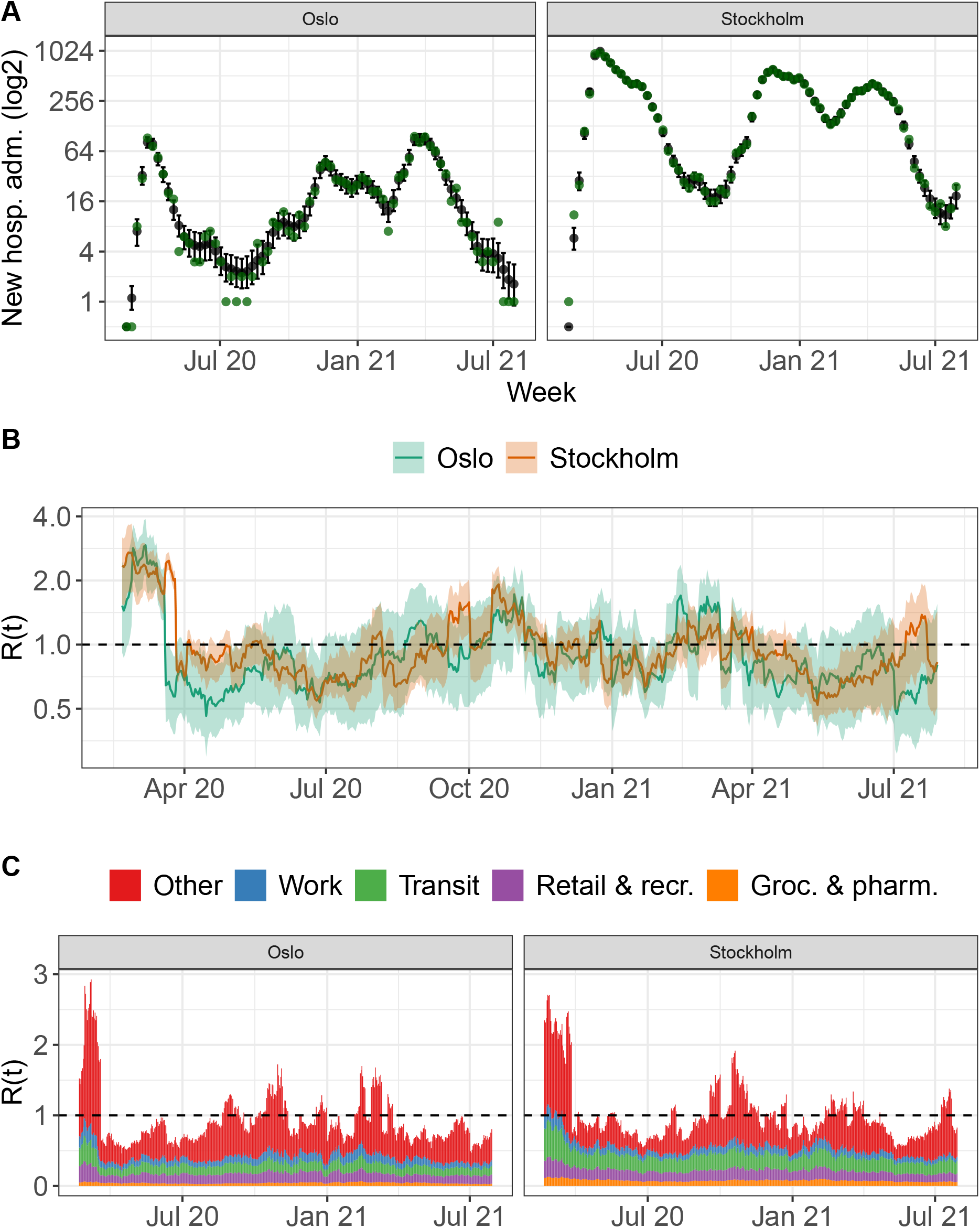
Model fit for two selected regions, Oslo and Stockholm. **Panel A** shows the weekly hospitalization counts: Oslo and Stockholm (green dots) as well as the model based expected hospitalization counts and associated 90%-credible interval. **Panel B** shows the estimated time-varying reproduction number, which is a function of the estimated 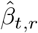 and corresponds to the expected number of secondary contacts per infected at each time-point and an associated 90%-credible interval. **Panel C** shows the model-based time-varying contribution of each transmission setting of the considered COVID-19 community mobility reports to these secondary infections.

In addition to changes in contact frequencies represented via the additive model for the GCMR, our model of SARS-CoV-2 transmissibility accounts for multiplicative changes in infection risk per contact associated with the Alpha and Delta VoCs and seasonal effects. We assume that these multiplicative effects are the same in all regions considered per model fit, and our estimates from the data of the Norwegian and Swedish regions are quite similar, as shown in Figure 2. We estimate that an increase in temperature of 20 degree Celsius (corresponding approximately to the difference between the average temperature of the coldest winter and warmest summer month) leads to a reduction in transmissibility by around 40%. Note that the median difference between the mean temperature in the warmest and coldest month of the year per region is 20.4 degrees in our observation period. For the Alpha VoC, we estimated an increase in transmissibility compared to the wildtype of around 20% based on data of the Norwegian regions but found no clear evidence for an increase in transmissibility in the Swedish data. For the Delta VoC, we estimated approximately a doubling of transmissibility compared to the wildtype in Norway and Sweden, albeit with substantial uncertainty.

**Fig. 2:**
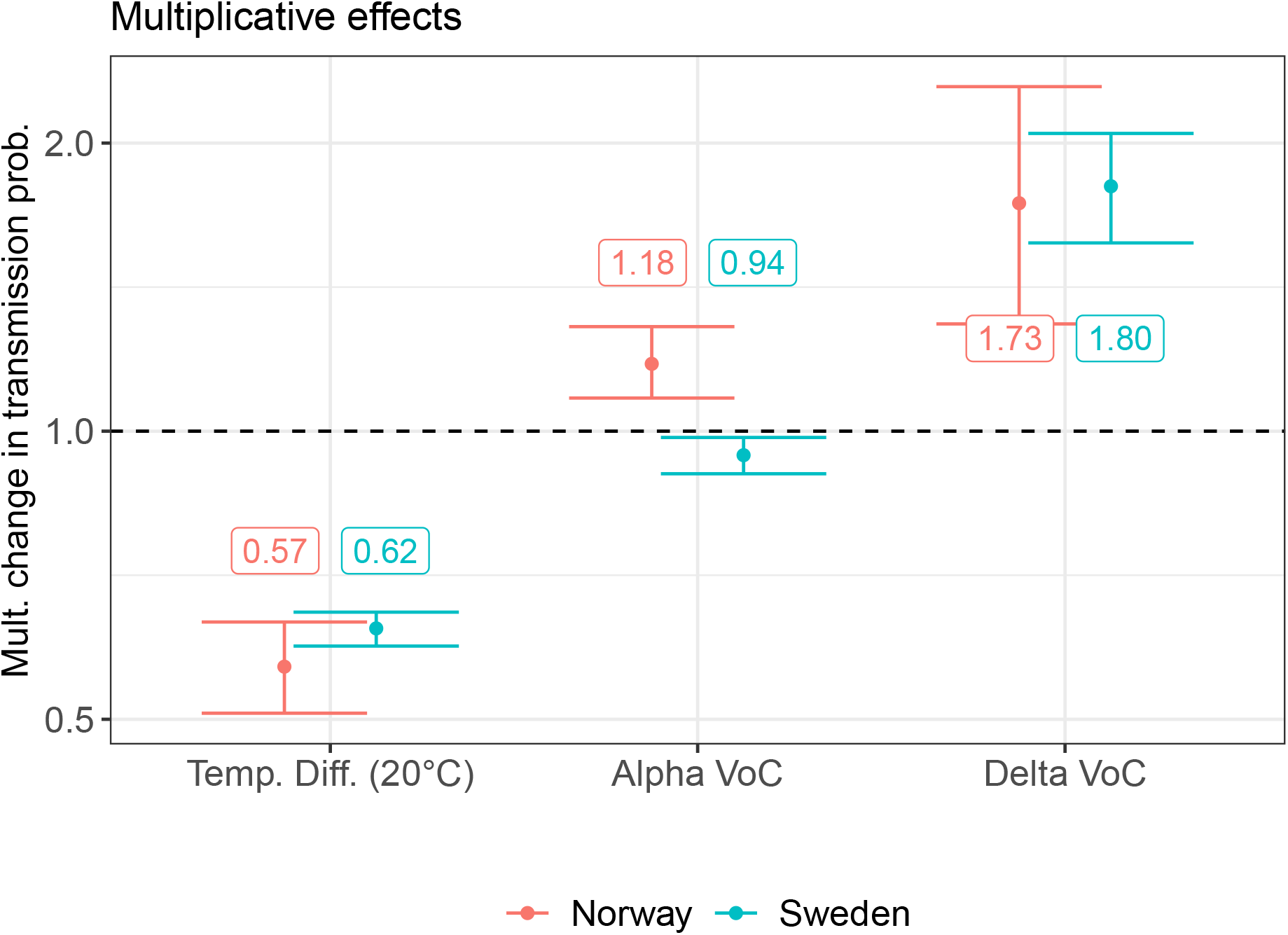
Multiplicative effects on transmission probability. Shown are the estimated posterior median and 90%-credible intervals for the multiplicative effects of VoC’s and a 20-degree temperature difference on the transmissibility for the Norwegian (red) and Swedish (blue) regions, respectively.

### 2.3 Model attributed 60% and 39% of all infections to contacts in four GCMR settings for Sweden and Norway

Based on our model, we derived the total share of infections that are attributed to activity/contacts in the GCMR categories per country or counties (Materials and Methods). Aggregated over the whole time period and all counties, our model allocated 60.0% (90%-CI: 57.1-62.5%) of all infections in Sweden and 38.7% (33.2 - 43.8%) of all infections in Norway to settings captured by the GCMRs. The remaining, unexplained, infectious contacts were accounted for by the region-specific weekly parameters *ν*_*week*(*t*),*r*_.

When investigating how much of the SARS-CoV-2 transmission dynamics was explained by the GCMRs in each county, we found rather large differences. The mean share of daily infectious contacts in settings captured by the GCMRs varied between 27.5 and 73.3%, the total share of infections assigned to the settings captured by the GCMRs was in a similar range, albeit slightly smaller for most regions (Fig. S25, Table S3).

The GCMRs explained a larger share of contacts in Swedish compared to Norwegian counties. We also observed a tendency towards a larger share of explained contacts for regions with higher population density. However, this correlation is rather weak and to a large extent driven by the capitals, Oslo and Stockholm, with high population density (Fig. S27).

### 2.4 In Norway and Sweden Transit stations and Workplaces are the most important GCMR settings of infection

Based on the estimated parameters *ϕ*_*k,r*_ from our model, contacts in settings captured by the Transit and Workplaces categories lead to most infections at the pre-pandemic baseline activity levels: the point estimate for the Transit setting was highest in 7 out of 11 Norwegian regions and in 9 out of 22 Swedish regions; in the remaining 4 regions in Norway and in 11 of the 12 remaining regions in Sweden, the Workplace setting was the most important setting (Supplementary table S5). Fig. 3 shows the population-weighted average of the relative share of secondary infections at baseline activity assigned to each of the four settings in Norway and Sweden, respectively. We estimate most infectious contacts in the Transit setting, followed by Workplaces, Retail & recreation, and lastly Grocery & pharmacies for Norway and Sweden. The posterior probability that the population-weighted mean of the Transit station setting is largest among the four settings is 0.55 for Norway and 0.68 for Sweden. Furthermore, we estimated a posterior probability of 0.50 and 0.62 that the settings Transit stations and Workplaces make up the settings with the largest coefficients among the six two-setting combinations.

**Fig. 3:**
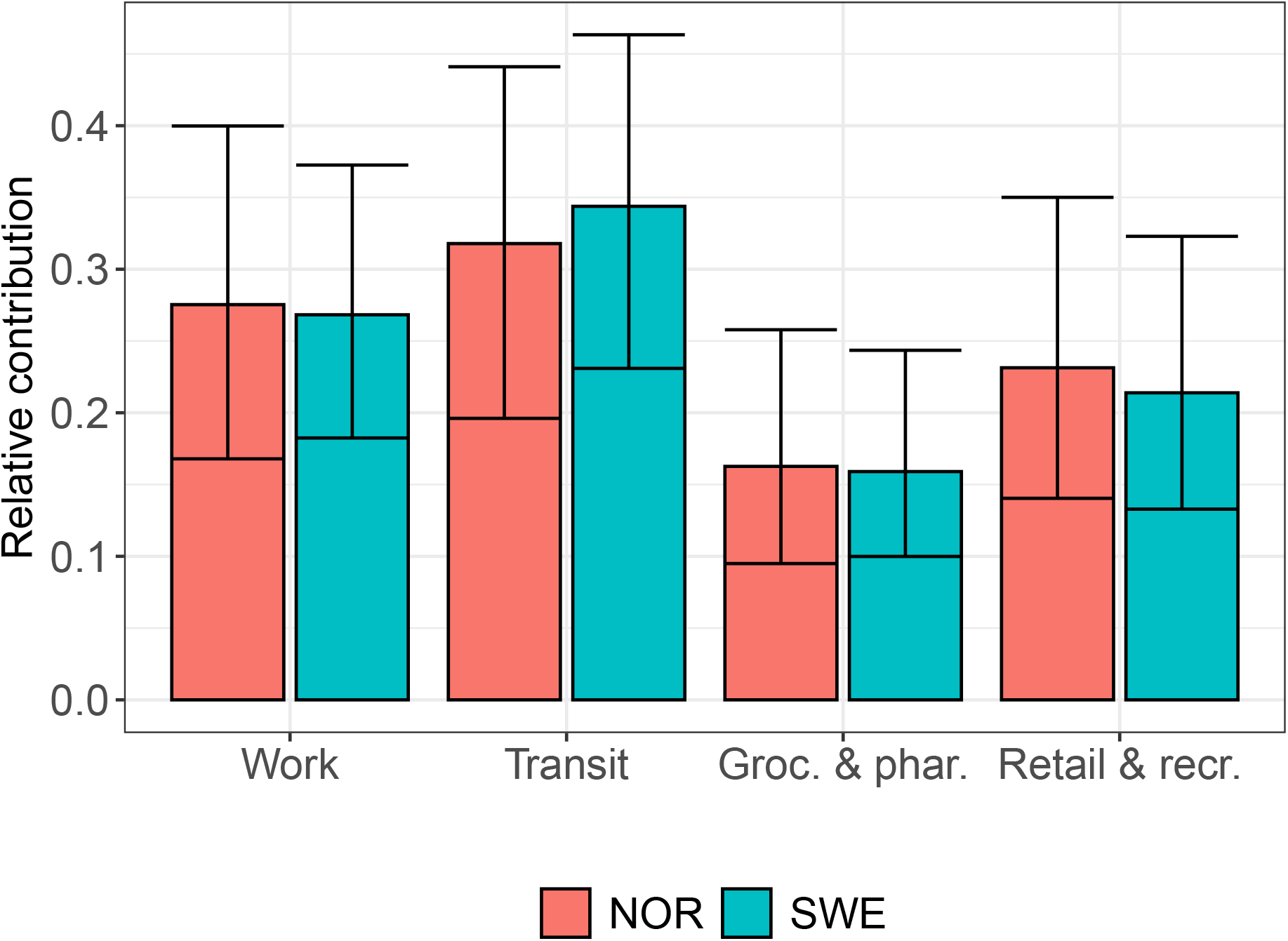
Population-weighted relative contribution of the community mobility reports at BL activity. Shown are the posterior median and 90%-credible intervals for the population-weighted relative contribution of each GCMR setting at baseline activity for Norway and Sweden.

### 2.5 Modeling German regions

Our proposed model to describe the relationship between GCMR and SARS-CoV-2 transmission rate, is not specific to Norway and Sweden, nor to the time period analyzed above. We estimated the model for two different regions from Germany, Bavaria and Berlin, in a shorter period (Feb 2020 - Dec 2020) based on publicly available data. For this period, no relevant virus variants or vaccination activity had to be considered (for more details, see Materials and Methods). Fig. 4 shows the model fit, estimated effective reproduction number and the contribution of the different GMR categories for Bavaria and Berlin. Compared to the results in the Norwegian and Swedish regions, the GCMR activity categories captured a relatively large share of infectious contacts (64.5% in Bavaria, 69.7% in Berlin on a daily average). During the observation period, 65.2% (90%-CI: 58.7-69.9%) of all infections in Berlin and Bavaria were allocated to contacts in settings captured by the GCMRs. The multiplicative effect of the temperature on the disease transmission in Germany had a similar order of magnitude compared to Sweden and Norway: a 20 degree increase in temperature reduced the model-based transmission probability per contact by factor 0.50 (90%-CI: 0.44-0.59). The largest difference compared to the results in Sweden and Norway was the model-based relative importance of the different GMR categories at baseline activity: In both German regions, and in particular Berlin, we estimated that activity in the Retail & recreation setting contributed most to infectious contacts, followed by the Transit and Workplace settings that where most important for Sweden and Norway (Fig. 5).

**Fig. 4:**
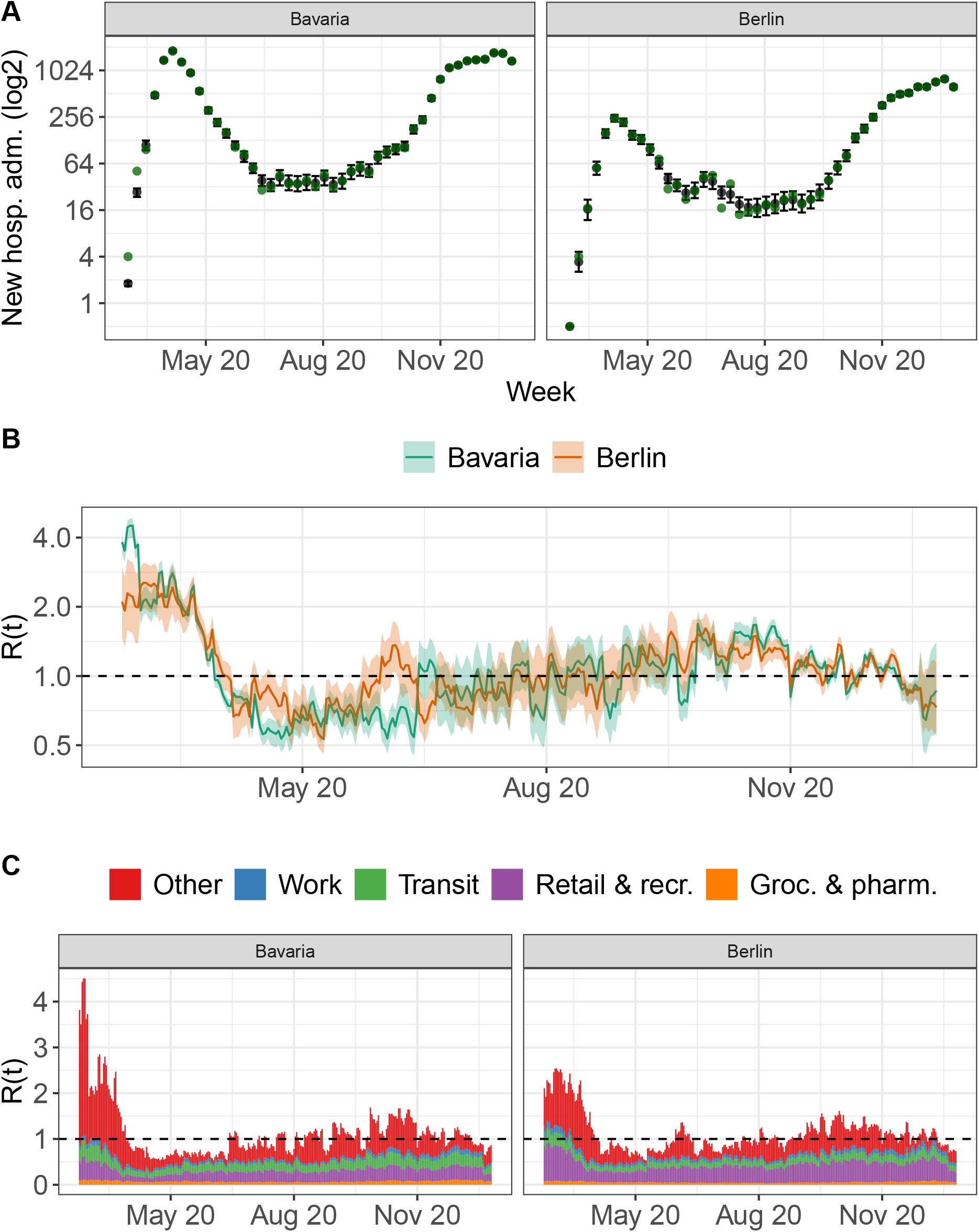
Model fit for the two German regions, Bavaria and Berlin. **Panel A** shows the weekly hospitalization counts: Bavaria and Berlin (green dots) as well as the model based expected hospitalization counts and associated 90%-credible interval. **Panel B** shows the estimated time-varying reproduction number, which is a function of the estimated 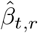 and corresponds to the expected number of secondary contacts per infected at each time-point and an associated 90%-credible interval. **Panel C** shows the model-based time-varying contribution of each transmission setting of the considered COVID-19 community mobility reports to these secondary infections.

**Fig. 5:**
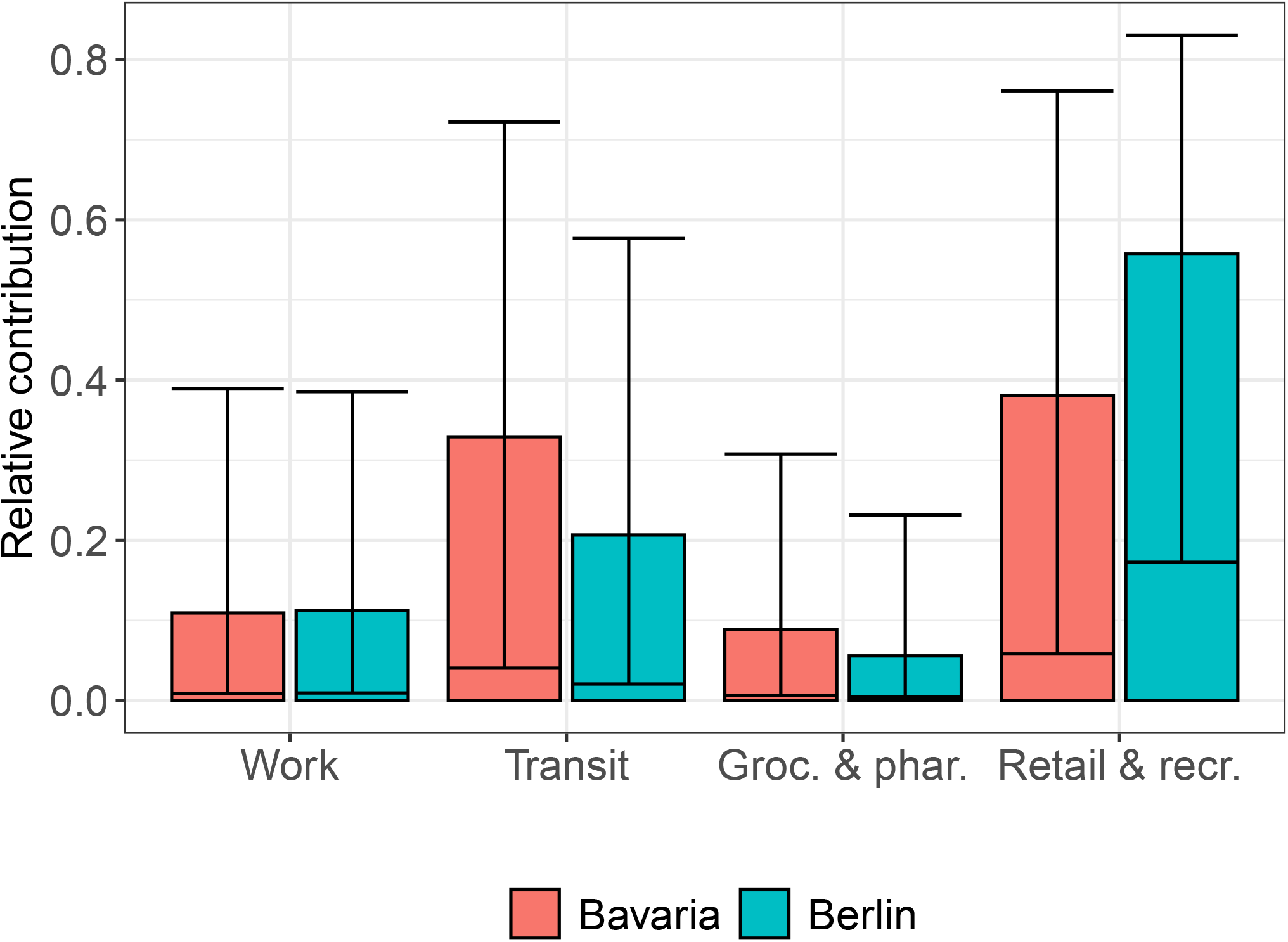
Relative contribution of the community mobility reports at BL activity, Bavaria and Berlin. Shown are the posterior median and 90%-credible intervals for the relative contribution of GCMR setting to the transmissibility at baseline activity, for the two analyzed German regions Bavaria and Berlin.

## 3 Discussion

We proposed a new model to quantify the contribution of contacts in different social settings to the spread of SARS-CoV-2, based on the Google Community Mobility Reports. In our state-space model for disease spread over time the transmissibility parameter was expressed as a function of the GCMRs in four social settings, as well as daily temperature and the proportion of different virus variants of concern. Importantly, the model was constructed to account for the non-standard definition of the GCMR indicators, so that each setting was modeled to have an additive contribution to the transmissibility. The rationale for this was that an increase/decrease in time spent in a specific social setting leads to an increase/decrease in transmission-relevant contacts in this setting, but does not affect transmissions in other settings. The temperature and virus variants on the other hand, were modeled to have a multiplicative effect on the transmissibility, thus affecting all types of transmission. Remaining changes in transmission dynamics were captured by a region- and week-specific random effect. This model allowed us to evaluate how well the GCMRs capture changes in disease dynamics during the SARS-CoV-2 pandemic, which settings of contacts appeared to be most relevant for transmission, as well as how disease transmission was related to temperature (as a proxy for seasonality) and virus variants. We applied the model to data from all Norwegian and Swedish regions and to two large German regions. This showed that the model is flexible and will be easy to apply to new situations.

The results of our analyses suggest that the GCMR data reflect changes in population behaviour that were directly related to the dynamics of the spread of SARS-CoV-2. Based on our model, for Sweden 60% of infections were attributed to social settings that were captured by the four GCMR on Workplaces, Transit stations, Groceries & Pharmacies, and Retail & Recreation, and 39% for Norway. The remaining infections cannot be explained by changes in population activity measured by the four GMCRs, the household setting most likely being one major additional infection setting. We observed not only differences in the explanatory power of the GCMRs between the two countries, but also relatively large differences between the individual counties.

We estimate that at regular, pre-pandemic activity-level contacts captured by the Workplace and the Transit station settings had the highest contribution to transmissibility among the four settings for both Norway and Sweden. However there were substantial changes in activity per setting over time. Therefore, these results do not necessarily imply that most infections happened, e.g., in public transport.

In our model, we accounted for seasonality and presence of virus variants by estimating multiplicative effects on the probability of infection upon contact between a susceptible and an infectious individual. For the VoCs, we also assumed an increase in hospitalization risk among infected by factor 1.9. We estimated a substantial change in transmissibility due to seasonality (transmissibility reduced by ∼ 40% in summer compared to winter) and increased transmissibility due to the the Delta VoC compared to the wildtype. In the Norwegian model, we also estimated a slight increase in transmissibility for the Alpha VoC. These results were in line with the existing literature and the temperature effect was estimated consistently for Sweden and Norway.

We also fitted the model to two German regions, Bavaria and Berlin, for the first 10 months of the pandemic. In these regions and time period, the GCMRs explained a slightly larger share of infections compared to our analyses for Sweden and particularly Norway. Note, however, the longer study period in the latter case. Just like for Norway and Sweden, the Transit station setting had a high contribution to transmission while activity at Groceries and pharmacies played lesser a role for transmission. The seasonality effect was estimated similarly for the German regions as in Norway and Sweden. However the analyses of the German regions found that activity in settings captured by the Retail & recreation setting played the largest role for transmission and contacts captured by the Workplace category had only a smaller contribution, which was interestingly opposite to Sweden and Norway. It would be interesting to study further in detail what might cause such differences, comparing the social behaviour, the organisation of labour, the mobility systems and the demographic profiles in these regions.

The proposed model has limitations: First, we assumed that changes in the considered GCMR settings were linearly related to the effective number of transmission-relevant contacts. This means for example that we assume that if GCMR-measured activity at workplaces is reduced by, say, 20%, then the number of transmission-relevant contacts in workplaces is reduced by 20% as well. Furthermore, we assumed that this linear relationship was constant over time, ignoring potential behavioural changes in specific settings, for example due to increased distancing between people. Both assumptions are rather strong, though we believe reasonable at a first order. Also, we assumed that effects of seasonality and virus variants change transmission risk in a multiplicative way in all social settings. One could argue that the effect of seasonality/temperature could be setting specific, the infection risk at workplaces might be changed differently as in Public transport. Our model assumes an all-or-nothing effect of vaccination and a maintained full protection of vaccination during the modeling period. A more realistic modeling of vaccination would be to implement a partial immunity upon vaccination and a gradual waning over time. However, our study period is possibly short enough so that this assumption does not introduce a major error and waning of vaccine-induced immunity did not play a large role. Also, we specified several aspects of the model (e.g., generation time, duration of infectiousness, increased hospitalization risk due to VoCs) based on consolidated literature. Wrong assumptions on these parameters might bias our results. A final concern is about the possible measurement errors of the GCMRs, which may not accurately enough capture changes in the amount of the social interactions between individuals in the specific categories of interest defined in the GCMRs. This may limit the causal interpretability of our results related to the (relative) importance of contacts in the different settings.

Overall, our analysis found a clear association between the publicly available GCMRs and SARS-CoV-2 infection dynamics in Norway, Sweden and two German regions. We were thus able to show that such data can also be very useful for a real-time assessment of the level of relevant population activity in times of an epidemic/pandemic. However, the peculiar definition and structure of GCMRs does not facilitate a straightforward utilization in mathematical models of disease transmission. For this, it requires rather strong assumptions, a complex model and estimation of many parameters. This might partly explain the variability of our results for the different regions as well. It is conceivable that more direct measurements of (absolute, mean) contact frequencies in different settings would have a bigger explanatory power with respect to actual transmission dynamics, and the real-time generation of such data would therefore be desirable for the future.

## 4 Materials and Methods

For Norway and Sweden, all computations were run for a period of about 17 months, from February 21, 2020 to July 29, 2021 (75 weeks). The spatial resolution of the model was by fylke in Norway and län in Sweden. In total 32 regions were included in the analysis, 11 in Norway and 21 in Sweden. Lower spatial resolution, i.e., municipality level, resulted in higher degree of missing GMCR data due to anonymity issues and in lack of hospitalization data.

### 4.1 Google community mobility reports (GCMR)

Google provides a freely available data source, the Google COVID-19 Community Mobility Reports, a daily, region specific, activity level in different social settings (2). These data are based on tracking Google users, who have “Location History” enabled on their mobile devices. Geographic locations are classified into six settings: Grocery and pharmacy, Retail and recreation, Transit stations, Parks, Workplaces, and Residential areas. For the first four settings, the GCMR index measures the percentage increase/decrease in the daily number of visits at locations classified to each setting, relative to a prepandemic baseline level. The baseline level, one for each day of the week, is the average number of visits at the locations and setting, measured over 5 consecutive weeks prepandemic, between Jan 3 and Feb 6, 2020. The daily number of visits in a setting is then divided by the baseline number in that setting of the corresponding weekday, and scaled so that 0 corresponds to the baseline level, 100 corresponds to a doubling in the number of visitors and, e.g., −50 corresponds to a reduction in visitors by 50%. The residential and workplace settings measure the length of stay (in minutes) during a 24 hour period rather than number of visits and are otherwise defined analogously. An unspecified, small amount of noise is added to the data before computation of the baseline values and the daily counts to meet anonymity requirements (3).

Let 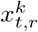 denote the Google covariate used in our model, at day *t* in region *r* in setting *k*. We have rescaled the data provided by Google, such that 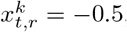, corresponds to a 50% reduction in activity level, 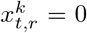 corresponds to the same activity level as at baseline and 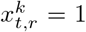 corresponds to a 100% increase in activity. Furthermore we used a (right-aligned) 7-day moving average of the data for our model to deal with the inherent weekday fluctuations in the GCMR data. Fig. S1A shows the mean GCMR and variation across regions for Norway and Sweden, for the different settings.

### 4.2 Temperature

Daily average temperature for Norway was acquired from the Norwegian Meteorological Institute Frost API, while the Swedish temperature data was acquired through the Open Data Meteorological Observations API provided by Swedish Meteorological and Hydrological Institute. For each region a weather station close to the most dense populated area was selected. The full list of stations selected is given in supplementary table S1.

### 4.3 Variants of concern data

The World Health Organization (WHO) declared the Alpha variant (pango lineage B.1.1.7) a Variant of Concern (VoC) on December 18, 2020. On May 11, 2021, WHO classified Delta (pango lineage B.1.617.2) as a VoC. Ideally we want to know the proportion of the true number of cases that are of the different variants. This is unknown because the true incidence in the population is unknown and not every case was sequenced. Instead, we approximate the true proportion by the proportion of the sequenced/screened cases, denoted 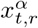 and 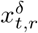 for the Alpha and Delta variants, respectively. The share of Alpha and Delta variants over time was only available on a national level in a weekly temporal resolution. The same time series were therefore used for all regions within a country. Supplementary figure S13 shows the data. For Norway, the data was acquired manually from the weekly Covid-19 reports of the National Institute of Public Health (9) and the Swedish data was acquired from the web pages of Public Health Agency of Sweden (10).

### 4.4 Vaccination data

Vaccination data is given as daily number of new individuals who have received the second vaccine dose, by region and by age group, denoted 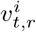 for day *t*, region *r* and age group *i*. See supplementary table S2 for age groups. Data for Norway was downloaded from the GitHub of The Norwegian Institute of Public Health (11) and data for Sweden was provided from the Public Health Agency of Sweden upon request. The vaccine data were not updated every day of the week. We therefore assume a linear increase in the cumulative number of vaccinated individuals at days with missing data. See Supplementary figure S15-S17.

### 4.5 Model for disease spread

We model the spread of disease over time in a region by a discrete time deterministic metapopulation model, an age-structured SIR model (see Supplementary figure S18). The population in region *r* at day *t* and age-group *i* is divided into susceptible individuals 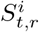, infectious individuals 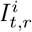, individuals recovered from infection 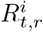 and vaccinated individuals *V*_*t,r*_. Transitions between the compartments are given by the following set of equations:

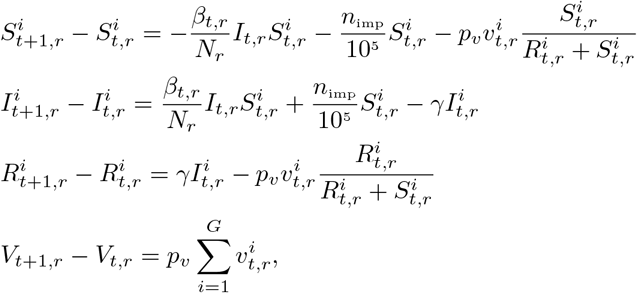

where *N*_*r*_ is the population size of region *r*. Infectious individuals at day *t*, 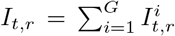, interact homogeneously with the susceptible individuals in each age group. The transmissibility parameter *β*_*t,r*_, is defined as the average number of infectious contacts an infected individual has at day *t* in a fully susceptible population. We model this parameter as a function of region specific covariates and associated parameters are estimated. In addition to the susceptible individuals infected by individuals from the infectious compartments, we assume that *n*_*imp*_ per 100 000 susceptible individuals in each age group are infected each day and moved to their respective infectious compartment, to mimic importation of cases. Infectious individuals in age group *i* are moved to the recovered naturally compartment 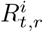 with a rate of 1*/γ* days.

The modeling period extends over a time period where vaccination was implemented. Age prioritized vaccination changes the age composition of the infected individuals, which then changes the risk of hospitalization. The likelihood of hospitalization at time *t* depends on the probability that a randomly selected susceptible individual belongs to a specific age group, hence, we need to keep track of the number of susceptible individuals in each age group. This motivates the age structure of the metapopulation model.

Vaccinated individuals are assumed to either become fully immune, hence not contributing to further transmission of the disease, or else, with probability *p*_*v*_ the vaccine has no effect. The daily number of new individuals with full immunity from vaccine, 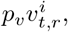, are removed from the susceptible and naturally recovered compartments, according to the relative size of 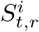 and 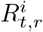. Immunity is assumed to remain over the study period.

### 4.6 Parameterisation of transmissibility parameter

We want to relate the transmissibility parameter *β*_*t,r*_ to the GCMR, temperature and variant covariates. The different GMCR settings are assumed to have an additive effect on the transmissibility, while the temperature and variant covariates are assumed to have a multiplicative effect.

The additive part of the transmissibility, 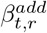, as a function of the GCMRs, is derived as follows. Assume that the average number of secondary infections of an infectious individual occur in *K* different settings, e.g. at work, public transport etc. For setting *k*, we define the average number of secondary infection at day *t* in region *r*, 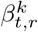, as the product between the average number of contacts in that setting, 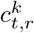, and the probability that a contact results in an infection, 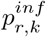. The number of contacts in each setting is unknown. By assuming a linear association between the GCMRs and the number of contacts in a setting, the number of secondary infections in setting *k* becomes

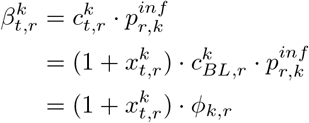

where 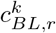 is the baseline level (pre-pandemic) number of contacts in setting *k* in region *r*. The parameter *ϕ*_*k,r*_ can be interpreted as the average daily secondary infections per infectious individual in region *r*, in setting *k* at the baseline activity level, i.e., pre-pandemic level when 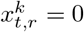.

The number of secondary infections caused by an infectious individual during a day is then the sum of secondary infections in the different settings. In addition to the *K* settings, we include daily secondary infections happening outside of the consiedered GCMR transmission settings by adding a weekly region specific parameter, *ν*_*week*(*t*),*r*_ (e.g. household transmission), assumed to be a priori i.i.d. between weeks and across regions based on a half-normal prior with region-specific standard deviation (cf. Implementation). The additive part of the transmissibility at day *t* in region *r* is then

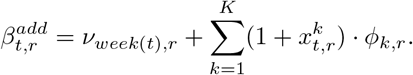

In our model fitting, we included *K* = 4 settings from the GCMR covariates: Grocery and pharmacy, Retail and recreation, Public transport and Workplace. We excluded the Park and Home settings as the assumption of a linear increase in infectious contacts with increasing activity seems less plausible for these settings.

Transmission dynamics are not only related to changes in contact frequencies. Seasonality and virus variants alter transmissiblity of a pathogen substantially. We use region-specific daily mean temperature as a proxy for seasonality and assume that the seasonality as well as the Alpha and Delta virus variants act multiplicatively on the transmissibility by changing the probability of infection upon contact. The multiplicative part of the transmissibility is modeled as

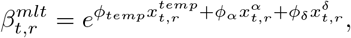

where 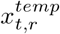 is the temperature difference at day *t* compared to the first day of the analysis in region *r*, and 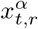 and 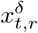 represent the fraction of individuals infected by the two variants of concern. The parameters *ϕ*_*temp*_, *ϕ*_*α*_ and *ϕ*_*δ*_ are assumed constant over region and in time. The multiplicative change in transmission probability for a one-degree increase in temperature is then 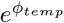, and 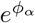 or 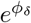 correspond to a multiplicative increase in transmissibility for the Alpha or Delta VoC compared to the wildtype. To characterize the total effect of seasonality we focus on the multiplicative change for a difference in temperature of 20 degrees, 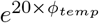, which corresponds to the median temperature difference between the average temperature in the warmest and coldest months of the year for the Norwegian and Swedish regions.

Combining the additive and multiplicative parts 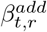 and 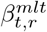, we get the final parameterisation of the transmissibility

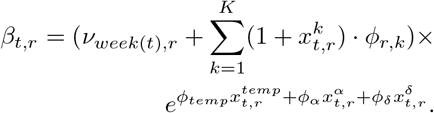

The parameters that we need to estimate in the above parameterization of *β*_*t,r*_ are *ν*_*week*(*t*),*r*_ (for 75 week and each region), *ϕ*_*r,k*_ (for each region and *K* = 4 behavior settings), *ϕ*_*temp*_, *ϕ*_*α*_ and *ϕ*_*δ*_.

### 4.7 Interpretation of model parameters and derived quantities

We focus on two aspects when interpreting the estimated parameters *ϕ*_*k,r*_ for the GCMRs. First, the proportion of variation in the additive part of transmissibility, which is explained by the linear model in combination with the observed change in the community mobility report data. For this purpose, we can quantify per region the mean share of mobility reports in 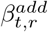 over time: 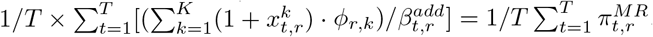. We refer to this quantity as the mean share of daily infectious contacts in settings captured by the community mobility reports. A similar summary statistic also takes into account the time-varying number of susceptible and infected individuals: 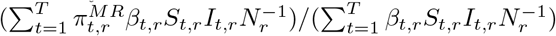. It represents the overall share of secondary infections in settings captured by the community mobility reports model over the whole observation period. A second aspect is related to which of the four considered community mobility reports appears to be most relevant for disease transmission. This can be addressed by investigating the relative size of the coefficients *ϕ*_*k,r*_ for the *K* = 4 categories: 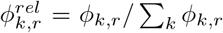. We perform posterior inference for this *ϕ*^*rel*^ per region and combine the region-specific estimates based on a population-weighted mean over all regions of a country.

### 4.8 Fitting the model

We fit the model to weekly number of new hospital admissions due to SARS-CoV-2 infection in each region. The number of hospital admission in week *w* is assumed to follow a Poisson distribution, with expectation dependent on daily new number of infected individuals, distribution of time between infection and hospitalization and risk of hospitalization given infection. Due to vaccination, the age distribution of the daily new infected individuals changes over time, which further changes the age-specific risk of hospitalization. For this reason, and the fact that hospitalization risk changes for different virus variants (21; 22), the modeled risk of hospitalization is time dependent.

### 4.9 Hospital incidence data

Hospital incidence data for Norway was acquired by request from Norwegian Institute of Public Health. The data consist of weekly number of new hospital admission, for individuals with Covid-19 infection as main cause of hospitalization, for each of the 11 Norwegian regions.

Swedish data on hospital incidence was obtained from Socialstyrelsen (18). They provide data on the weekly hospital incidence on a national level, as well as the number of currently treated individuals (occupied beds) on the county-level. County-level hospital incidence was not directly available. To approximate weekly hospital-incidence on the county-level, we distributed the national incidence to the counties based on the relative share of all individuals treated per county within the respective week.

### 4.10 Likelihood of hospitalizations

The number of new hospitalizations, *H*_*w,r*_, in week *w* in region *r* was modeled by a Poisson distribution,

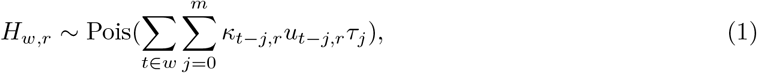

where *κ*_*t,r*_ denotes the probability of being hospitalized if infected at day *t* and belonging to region *r*; *m* is the maximum number of days from infection to hospitalization and *τ*_*j*_ denotes the probability of going to hospital *j* days after infection, given that an infected individual goes to hospital. The quantity *u*_*t,r*_ is the number of new infections at day *t* in region *r* in our model, given by

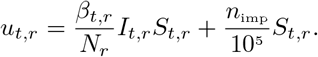

If all infected individuals were hospitalized, then the expected number of individuals sent to hospital at day *t* will be the sum, over the preceding *m* days, of the number of new infections *j* days before day *t*, times the probability *τ*_*j*_ that those new infections are hospitalized after *j* days, i.e., at day *t*. Not all infected individuals need hospitalization, and we therefore multiply by *κ*_*t*−*j,r*_, the probability that an individual infected at day *t* − *j* goes to hospital at day *t* (age-differences taken into account as explained below). The expected new hospitalizations in week *w* is then the sum over all days *t* in week *w*.

### 4.11 Distribution of time between infection and hospitalization, *τ*_*j*_

The probability of being hospitalized *j* days after infection, given that an infected individual gets hospitalized, is denoted *τ*_*j*_. The time of infection in our context is the time from the beginning of the infectious period, i.e. when an individual is moved to the infectious compartment. The time from infection to hospitalization consists of a pre-symptomatic period (time from infectiousness starts to symptom onset), followed by the period from symptom onset to hospital admission.

The situational awareness and forecasting reports (1), Week 51, 29 December 2021, assumes that the pre-symptomatic period is exponentially distributed with mean 2 days, i.e., *Exp*(1*/*2). Norwegian data on time from symptom onset to hospitalization, provided by FHI on request, fits well with a negative binomial distribution, with mean and overdispersion parameter changing with time. We assume these parameters are constant in time, and therefore resample data from the negative binomial for each time period, and from this new data set estimate the mean (7.93) and overdispersion parameter (6.21) of a negative binomial distribution.

We assume that the length of the pre-symptomatic period and the symptomatic period are independent, and hence, *τ*_*j*_ is expressed as the convolution between the exponential distribution of the pre-symptomatic period and the negative binomial distribution of the time from symptom onset to hospitalization.

### 4.12 Probability of hospitalization given infection, *κ*_*t,r*_

The probability of being hospitalized if SARS-CoV-2 infected is highly age dependent. The age distribution of the infected individuals in a region changes with age prioritized vaccination. The probability of hospitalization is also affected by the circulating virus variants. In the above likelihood of hospitalizations, the region specific parameter *κ*_*t,r*_ is time dependent, but not age specific. We therefore need the time variation of the parameter to take into account both the changing age distribution of the infected individuals and the changing proportion of virus variants.

Let 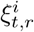 be the probability that an individual in region *r* infected at day *t* goes to hospital, given that the individual belongs to age group *i*. The probability that an individual infected at time *t* in region *r* is admitted to hospital is expressed as

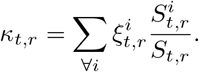

where 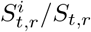 is the fraction of the susceptibles being in age group *i* in region *r* at day *t*. We have used values for 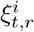 for the “wild-type” variant as given in the FHI modeling reports (1). These probabilities are built on work by Salje et al. (17) and adjusted to fit a Norwegian population. However, vaccine data both for Norway and Sweden have different age groups, so we need to transform these probabilities to fit other age groups. A simple approach to this is to fit the exponential function *y* = *e*^(*a*+*bx*)^, where *x* denotes age, to the Norwegian specific probabilities. The estimated coefficients were *a* = −2.52 and *b* = 0.06. Probabilities for the new age groups were then inferred by choosing the fitted value *y* that corresponds to the middle age of each age group. See Supplementary figure S14 and Supplementary table S2 for age specific hospitalization risk for Norway and Sweden.

These risks of going to hospital for a given virus variant and age group is assumed constant throughout the pandemic, however, they change for different virus variants. The proportion of the different variant among the infectious individuals change over time, and hence do the risk of hospitalization. The risk of going to hospital if infected with the Alpha variant is estimated to be 1.9 times higher than if infected by the Wildtype (21; 22). The same holds for the Delta variant. This increase in hospitalization risk is assumed to be the same for all age groups. The overall risk of going to hospital, given infection, in terms of the proportion of the Alpha variant 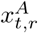 and Delta variant 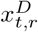 is then

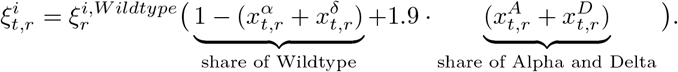

We only have access to variant data on a national level and therefore use the same data for all region within a country, despite the variants have probably spread differently in the different regions (9).

### 4.13 Implementation

The metapopulation and data model was implemented in Stan using the `cmdstanr` R-package to facilitate parameter estimation via Hamiltonian Monte Carlo. For a model fit to *n*^*reg*^ regions (11 for Norway, 21 for Sweden, and 2 for Germany) with 4 GCMR categories, 3 multiplicative factors on transmissibility (Temperature, and change in tranmissibility for Alpha and Delta VoC) in a period of *n*^*week*^ weeks (75 for Norway/Sweden, 44 for Germany) we estimated

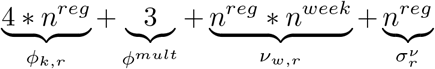

parameters and hyper-parameters. We used the following Normal and (truncated) Half-normal priors:

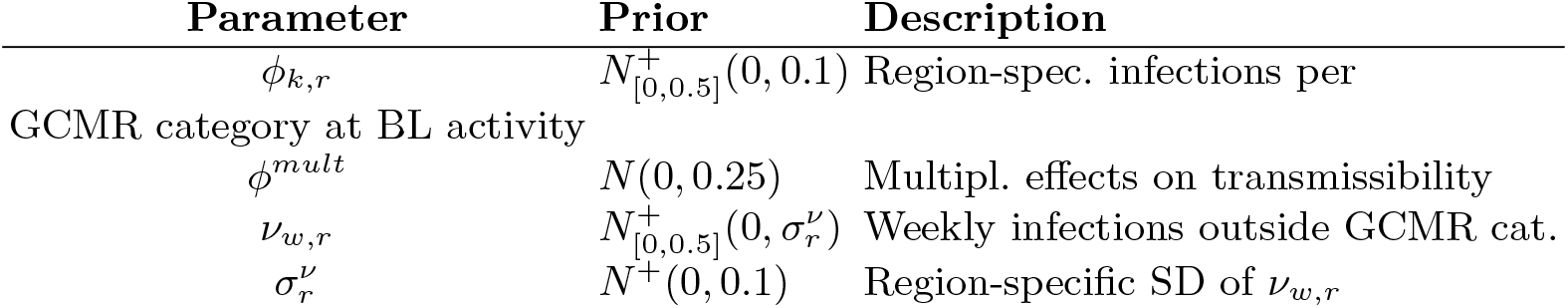

| Parameter | Prior | Description |
| --- | --- | --- |
| $\phi_{k,r}$ | $N_{[0,0.5]}^+(0, 0.1)$ | Region-spec. infections per |
| GCMR category at BL activity |  |  |
| $\phi^{mult}$ | $N(0, 0.25)$ | Multipl. effects on transmissibility |
| $\nu_{w,r}$ | $N_{[0,0.5]}^+(0, \sigma_r^\nu)$ | Weekly infections outside GCMR cat. |
| $\sigma_r^\nu$ | $N^+(0, 0.1)$ | Region-specific SD of $\nu_{w,r}$ |

The data model for the region-specific weekly hospitalization counts consisted of a Poisson Likelihood as described above. The model was fitted using 4 independent chains with with 1000 samples (200 warmup) each and convergence was monitored based on standard diagnostics (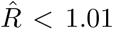 for all parameters and visual investigation of trace plots). Source code including the Stan-model as well as the full analysis pipeline for the public data from Germany (based on the `targets` R-package) are provided in a Github repository https://github.com/FelixGuenther/nordic_behavior_public.

### 4.14 Analysis for German regions

For fitting our model to two German regions we used 44 weekly data points on the reported 7-day hospitalization incidence provided by the Robert Koch-Institute from 1 March, 2021 to 27 December, 2022 for the federal states of Berlin and Bavaria (16). These measurements contain all reported hospital cases from the last 7 days. We collected the publicly available GCMRs and obtained temperature data from the openly available database of Deutscher Wetterdienst. As this analysis was focused on the year 2021 only, no data on vaccination or VoCs was required. Otherwise, model specification and fitting was performed as in the analysis for Norwegian and Swedish counties with one exception: as the population size of the German federal states is substantially larger compared to the Nordic counties we reduced the daily importation of infectious individuals from 0.5/100,000 individuals to 0.1/100,000 to avoid an unrealistically high assumption on (absolute) imported cases in particular during low-incidence periods.

## Data Availability

All data produced in the present study are available upon reasonable request to the authors.
Code and data for a central part of the analysis are available at:
https://github.com/FelixGuenther/nordic_behavior_public/
Publicly available data on COVID-19 hospitalization counts were obtained from
NOR:
https://www.fhi.no/en/id/corona/coronavirus/daily-reports/daily-reports-COVID19/Data
SWE:
https://www.socialstyrelsen.se/statistik-och-data/statistik/statistik-om-covid-19/
GER:
https://github.com/robert-koch-institut/COVID-19-Hospitalisierungen_in_Deutschland/

https://github.com/FelixGuenther/nordic_behavior_public/

## Notes

### Competing Interest Statement

The authors have declared no competing interest.

### Funding Statement

This work was supported by the Nordforsk project 105572. AF and HKB were also supported by BigInsight, the Norwegian Research Council project number 237718. AF was also supported by Integreat, the Norwegian Research Council project number 332645.

### Author Declarations

The study used publicly available data on the number the number of Covid-19 hospitalizations, vaccination numbers, and share of virus variants as provided by the Public health agencies of the considered countries (Sweden, Norway, and Germany). The finest level of aggregation was weekly (cumulative) numbers at county/federal state level. Data was obtained from the following web ressources: https://www.fhi.no/en/id/corona/coronavirus/daily-reports/daily-reports-COVID19/Data https://www.socialstyrelsen.se/statistik-och-data/statistik/statistik-om-covid-19/ https://github.com/robert-koch-institut/COVID-19-Hospitalisierungen_in_Deutschland/

