## SupplementaryMaterial for "Quantifying the impact of social activities on SARS-CoV-2 transmission using Google mobility reports"

### Supplementary Text 1 - Substantial changes of population behaviour between February 2020 and July 2021

The COVID-19 community mobility reports provided by Google (1) represent a relative percentage change in population activity in six different categories: Workplaces, Transit stations, Groceries and pharmacies, Retail and recreation, Residential areas and Parks. During our study period from February 21, 2020 to July 29, 2021, the mobility reports show substantial changes in population behaviour in Norway and Sweden (Fig. 1A). After an increasing number of SARS-CoV-2 cases were reported in Sweden and Norway in late February and early March 2020, initially mainly related to travel but then quickly showing clear signs of spread of the virus in the population, the measured activity level at workplaces, transit stations, as well as in areas for retail and recreation decreased substantially from mid-March. Time spent in residential areas, on the other hand, increased sharply during the same period. After the first pandemic wave, the activity level in the different categories gradually approached the pre-pandemic level again; in the summer and early autumn of 2020, the activity level in transport stations and residential areas was similar to the pre-pandemic level, and in grocery and pharmacy shops as well as in the retail and recreation sectors it was even slightly increased in some cases. In the workplace category, on the other hand, the activity level in the summer was again substantially reduced compared to the pre-pandemic baseline level in January and February 2020, presumably related to the seasonal effect of widespread holidays and working from home in the Nordic countries during the summer. In the late autumn and winter of 2020/2021, general activity was again significantly reduced, especially at workplaces, transit stations and retail and recreation, and time spent in residential areas was significantly increased. This trend continued - albeit less strongly - into spring and early summer 2021. In addition to behavioural changes related to the pandemic response (voluntary and recommended activity and contact reductions, as well as restrictions and lockdowns), the data also show clear seasonal trends, as well as changes attributable to (public) holidays. For example, there are clear behavioural changes around Easter, Midsummer, Christmas and New Year's eve and - especially in the workplaces and parks category - the summer months compared to the rest of the year.

The six different categories of the COVID-19 community mobility reports capture different settings of population activity, nevertheless they show (partly) strong correlation over time. The Retail and recreation category and Grocery and pharmacy categories show the strongest positive correlation, while there is also substantial correlation with changes in activity at Transit stations. Time spent in residential areas is negatively correlated with all other categories (Fig. 1B). Comparing the average behavioural changes in the Norwegian and Swedish regions, we find that the overall patterns of activity changes are similar in both countries, although they are often more pronounced in Norway (Fig. 1A).

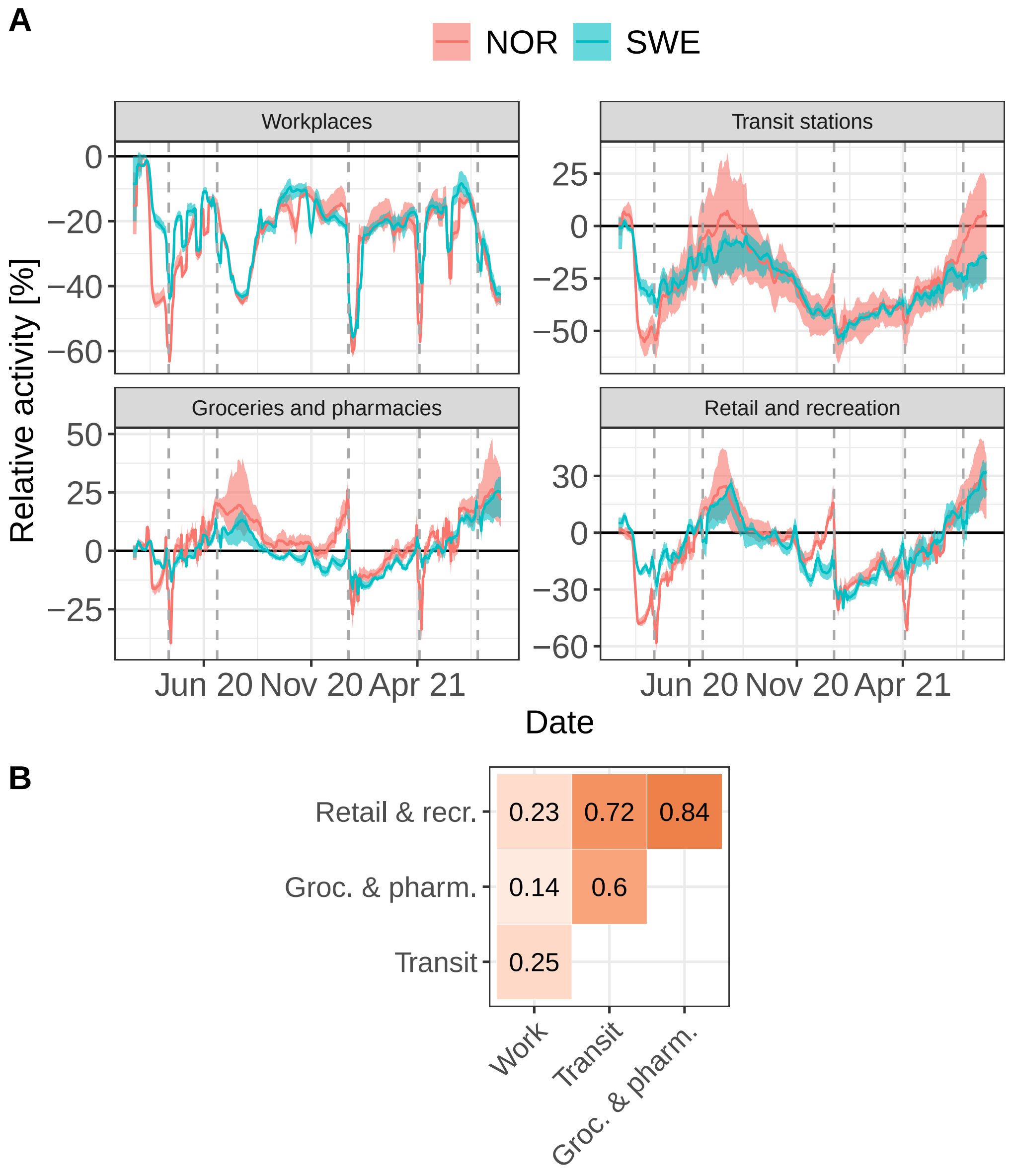


Fig. 1: **Descriptive visualisation of the Google COVID-19 community mobility reports for Norway and Sweden from February 21, 2020 until July 29, 2021. Panel A** shows the the average change in activity compared to baseline over all regions in Norway and Sweden, respectively. The ribbons represent 25%- and 75%-percent quantiles of the different regions. In each region, we smoothed the time-series based on a right-aligned 7-day moving average before calculating country-specific mean and quantiles. Dotted grey lines show main public holidays (East, Midsummer and Christmas). **Panel B** shows the pairwise Spearman correlation coefficient of the different (smoothed) activity categories over all regions.

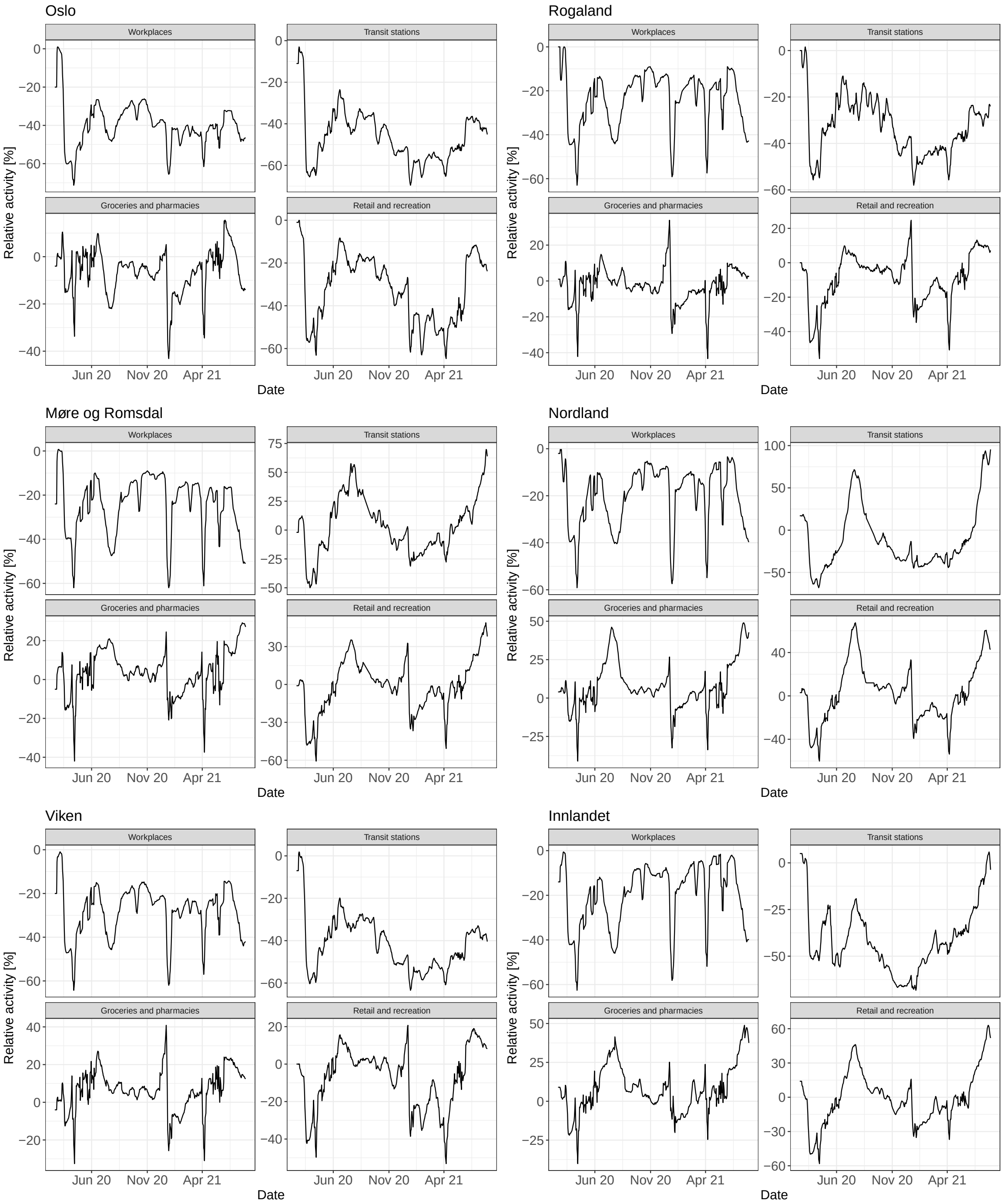


Fig. 2: **Visualisation of the Google COVID-19 community mobility reports for Norway from February 21, 2020 until July 29, 2021.** The figure shows the activity compared to baseline 6 of 11 regions in Norway. The lines are the smoothed time-series based on a right-aligned 7-day moving average.

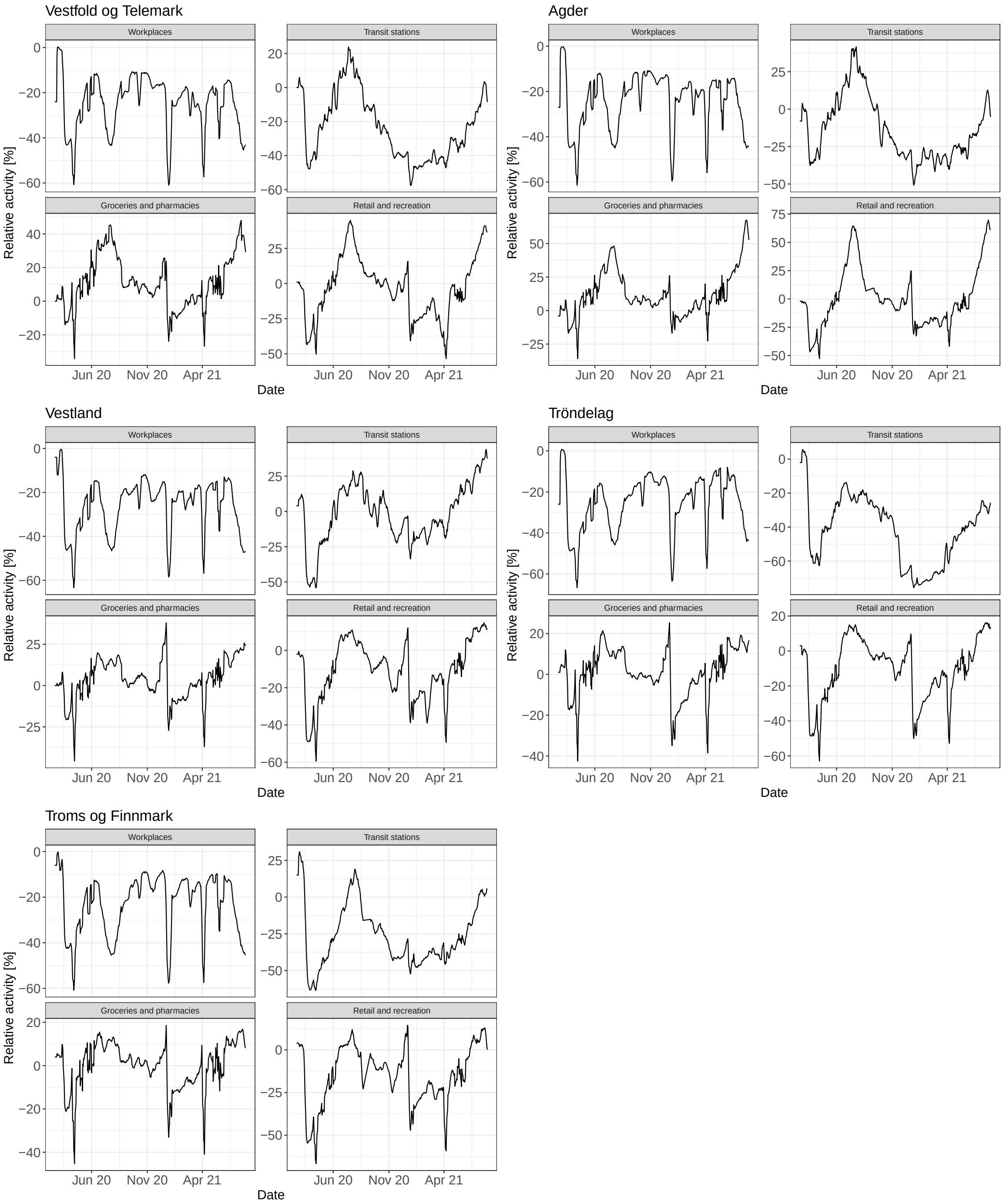


Fig. 3: **Visualisation of the Google COVID-19 community mobility reports for Norway from February 21, 2020 until July 29, 2021.** The figure shows the activity compared to baseline for 5 of 11 region in Norway. The lines are the smoothed time-series based on a right-aligned 7-day moving average.

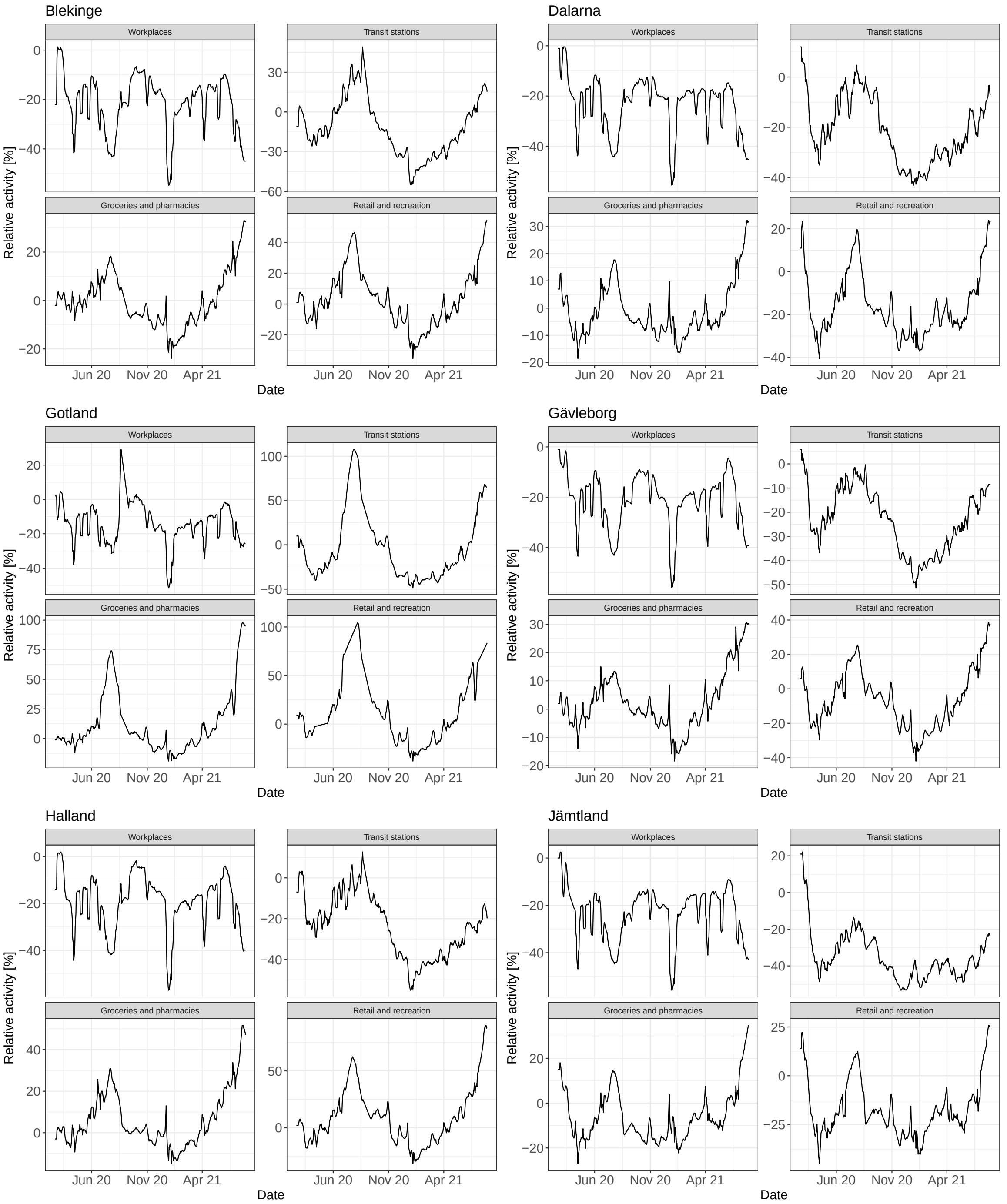


Fig. 4: **Visualisation of the Google COVID-19 community mobility reports for Norway from February 21, 2020 until July 29, 2021.** The figure shows the activity compared to baseline for 6 of 21 regions in Sweden. The lines are the smoothed time-series based on a right-aligned 7-day moving average.

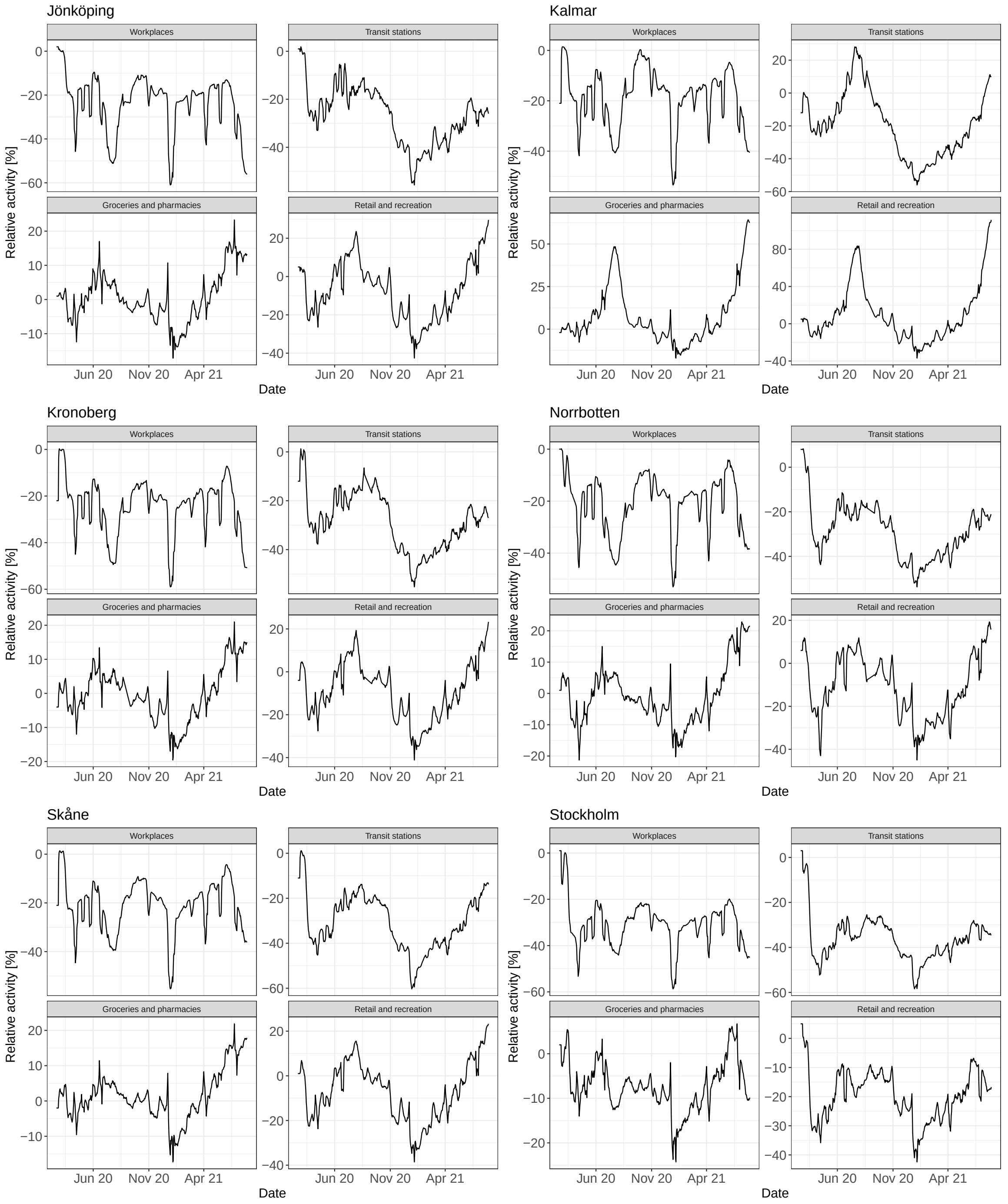


Fig. 5: **Visualisation of the Google COVID-19 community mobility reports for Norway from February 21, 2020 until July 29, 2021.** The figure shows the activity compared to baseline for 6 of 21 regions in Sweden. The lines are the smoothed time-series based on a right-aligned 7-day moving average.

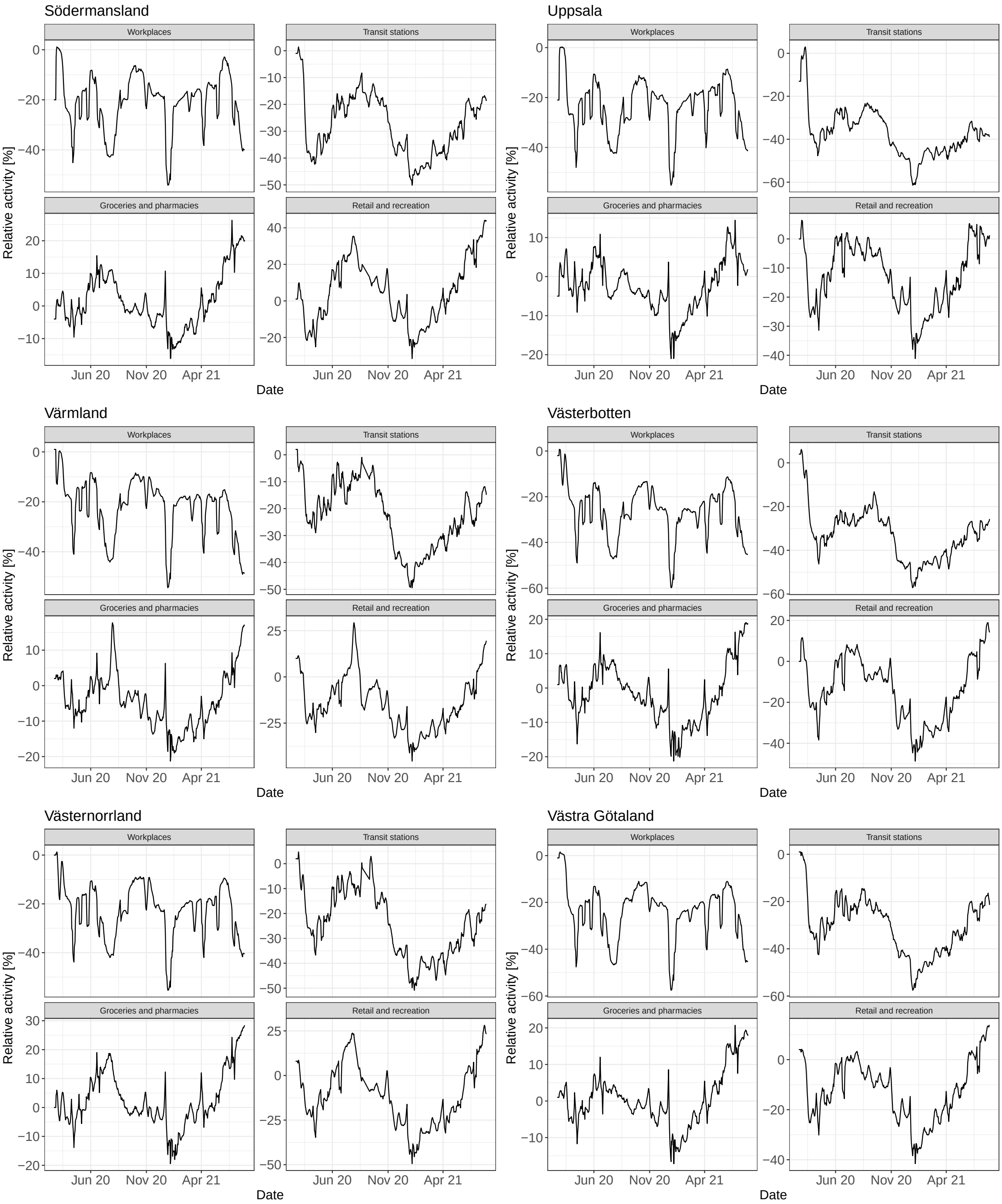


Fig. 6: **Visualisation of the Google COVID-19 community mobility reports for Norway from February 21, 2020 until July 29, 2021.** The figure shows the activity compared to baseline for 6 of 21 regions in Sweden. The lines are the smoothed time-series based on a right-aligned 7-day moving average.

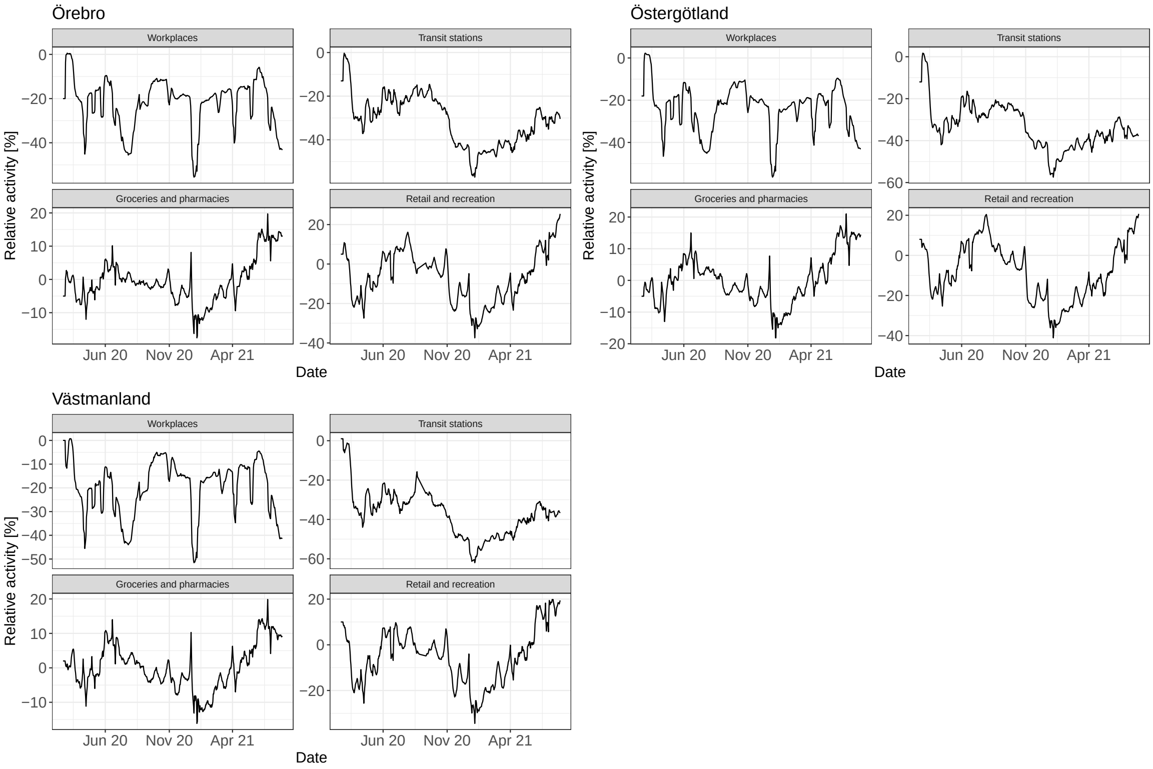


Fig. 7: **Visualisation of the Google COVID-19 community mobility reports for Norway from February 21, 2020 until July 29, 2021.** The figure shows the activity compared to baseline for 3 of 21 regions in Sweden. The lines are the smoothed time-series based on a right-aligned 7-day moving average.

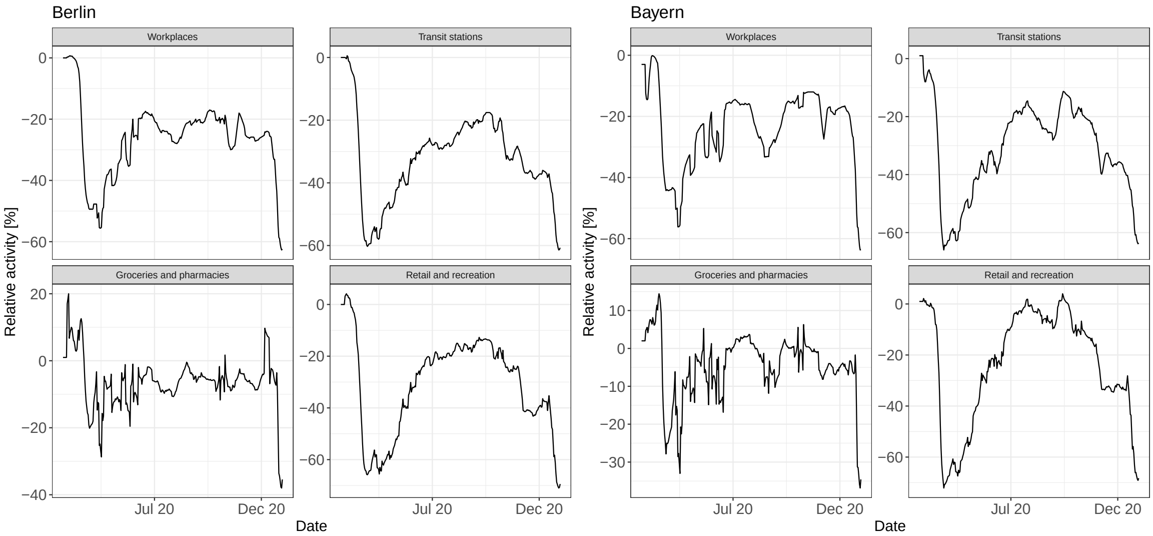


Fig. 8: **Visualisation of the Google COVID-19 community mobility reports for the German regions Bayern and Berlin, from February 21, 2020 until December 31, 2020.** The figure shows the activity compared to baseline. The lines are the smoothed time-series based on a right-aligned 7-day moving average.

### Temperature covariate

Table 1: Weather stations selected for each region in Sweden and Norway

| Country | Region | Station id |
|---|---|---|
| Norway | Oslo | SN18700 |
| Norway | Rogaland | SN44560 |
| Norway | Møre og Romsdal | SN60875 |
| Norway | Nordland | SN82290 |
| Norway | Viken | SN17850 |
| Norway | Innlandet | SN12680 |
| Norway | Vestfold og Telemark | SN30255 |
| Norway | Agder | SN39040 |
| Norway | Vestland | SN50540 |
| Norway | Trøndelag | SN68860 |
| Norway | Troms og Finnmark | SN90450 |
| Sweden | Blekinge | 65090 |
| Sweden | Dalarna | 104580 |
| Sweden | Gotland | 78400 |
| Sweden | Gävleborg | 107420 |
| Sweden | Halland | 63590 |
| Sweden | Jämtland | 134410 |
| Sweden | Jönköping | 74460 |
| Sweden | Kalmar | 66420 |
| Sweden | Kronoberg | 64510 |
| Sweden | Norrbotten | 162860 |
| Sweden | Skåne | 52350 |
| Sweden | Stockholm | 97200 |
| Sweden | Södermansland | 96190 |
| Sweden | Uppsala | 97510 |
| Sweden | Värmland | 93220 |
| Sweden | Västerbotten | 140480 |
| Sweden | Västernorrland | 127310 |
| Sweden | Västra Götaland | 71420 |
| Sweden | Örebro | 95130 |
| Sweden | Östergötland | 85240 |
| Sweden | Västmanland | 96560 |

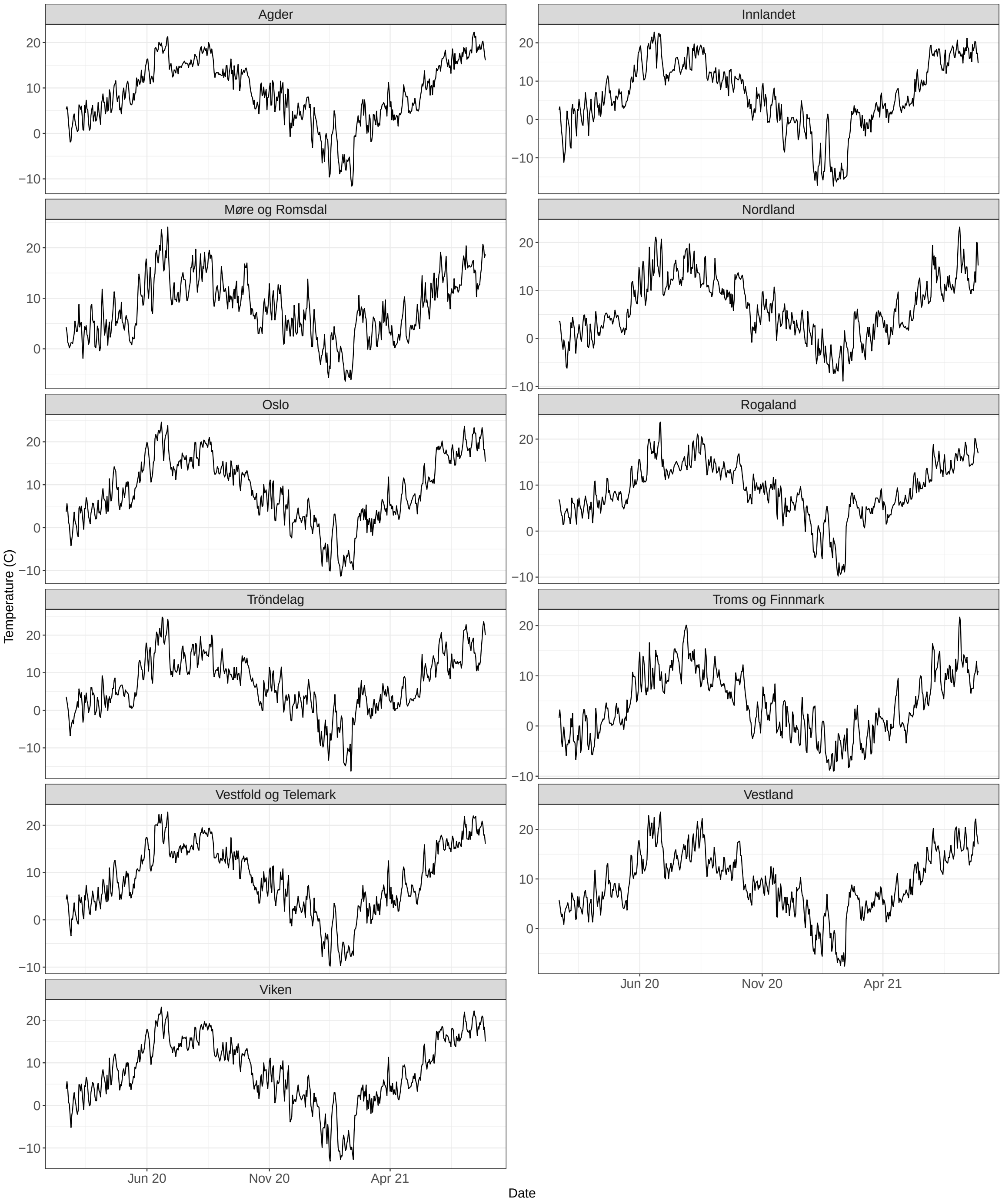


Fig. 9: **Temperature data by region for Norway from February 21 , 2020 until July 29, 2021.**

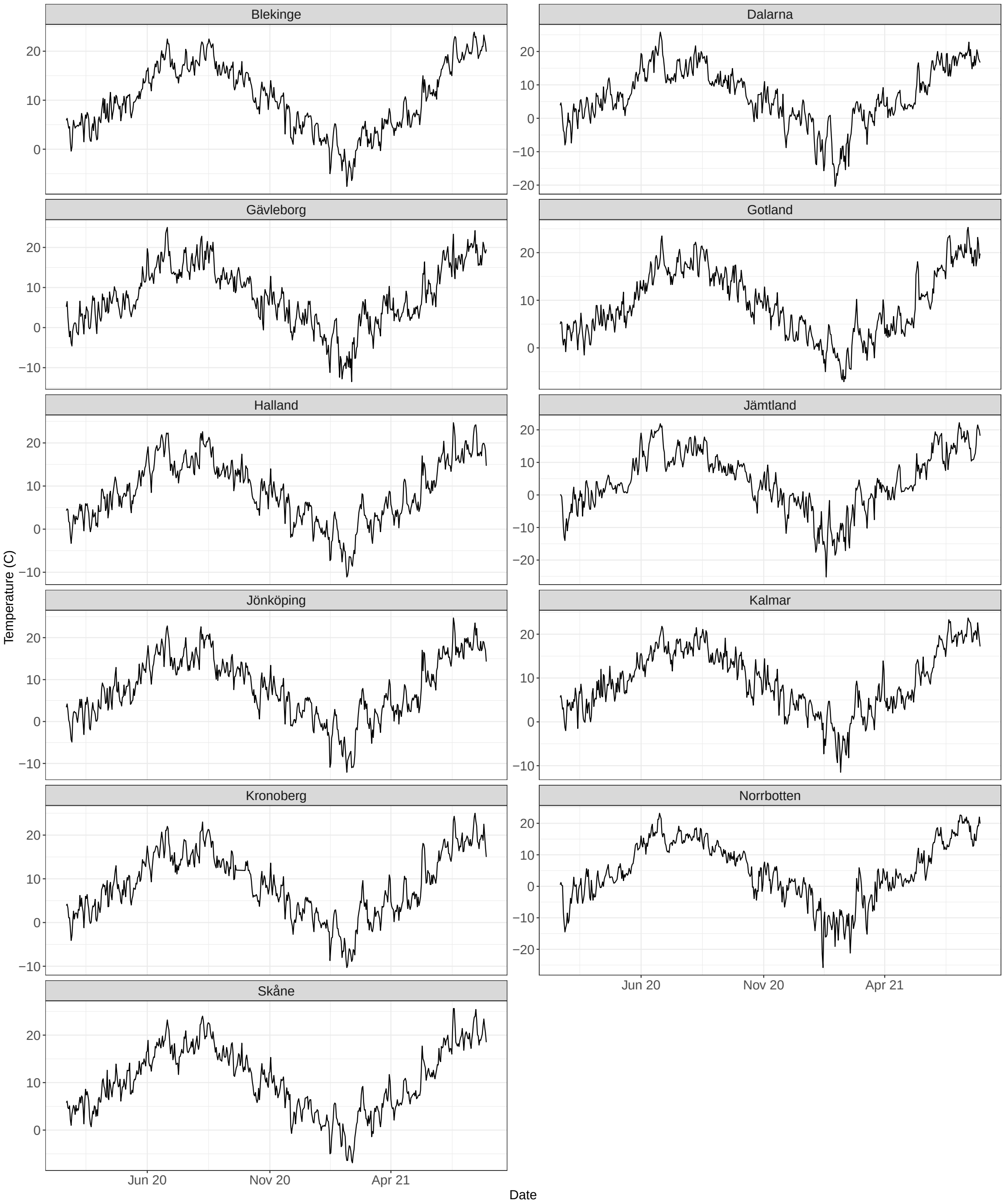


Fig. 10: **Temperature data by region for 11 regionsin Sweden, from February 21 , 2020 until July 29, 2021.**

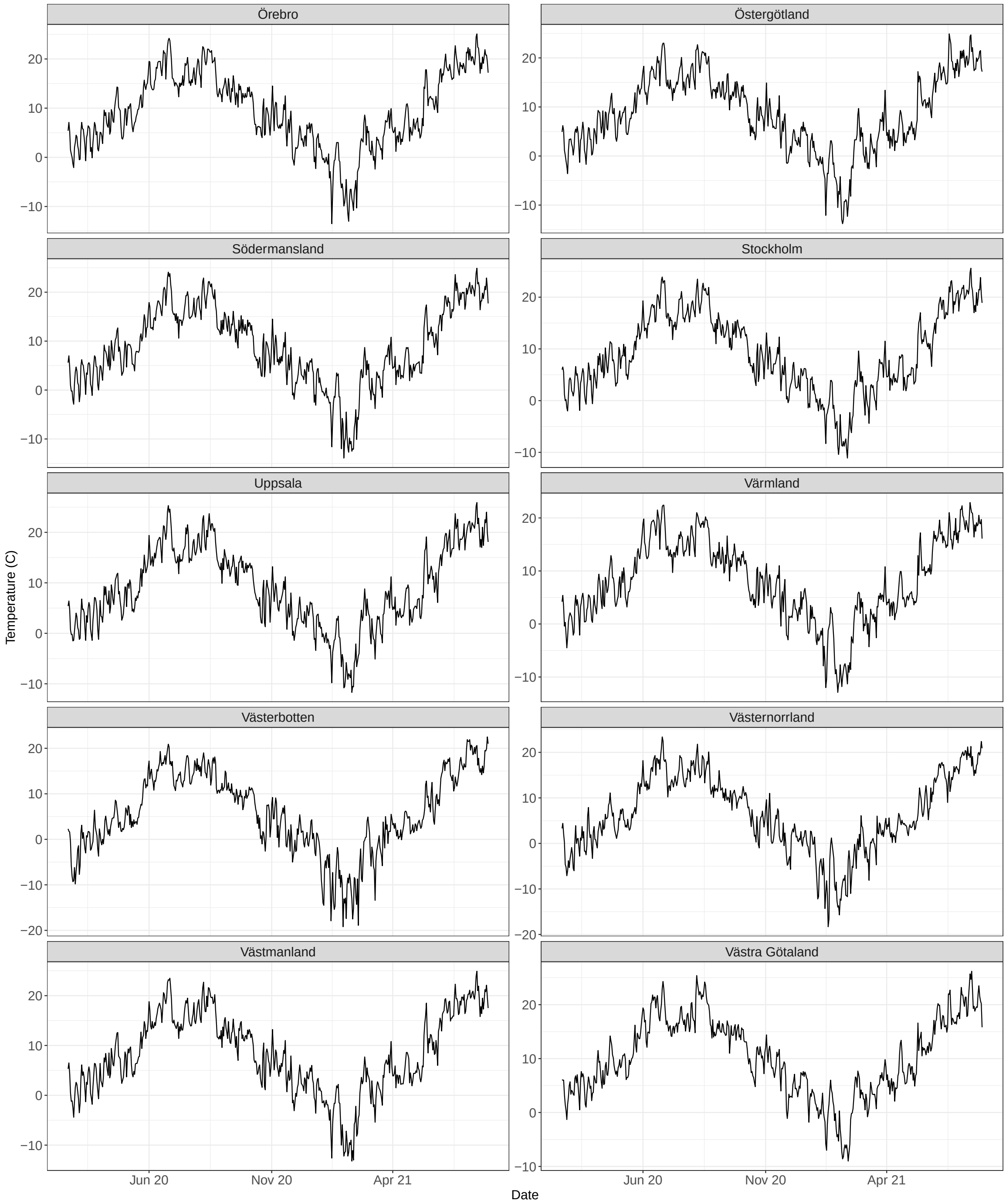


Fig. 11: **Temperature data by region for 10 regions in Sweden, from February 21 , 2020 until July 29, 2021.**

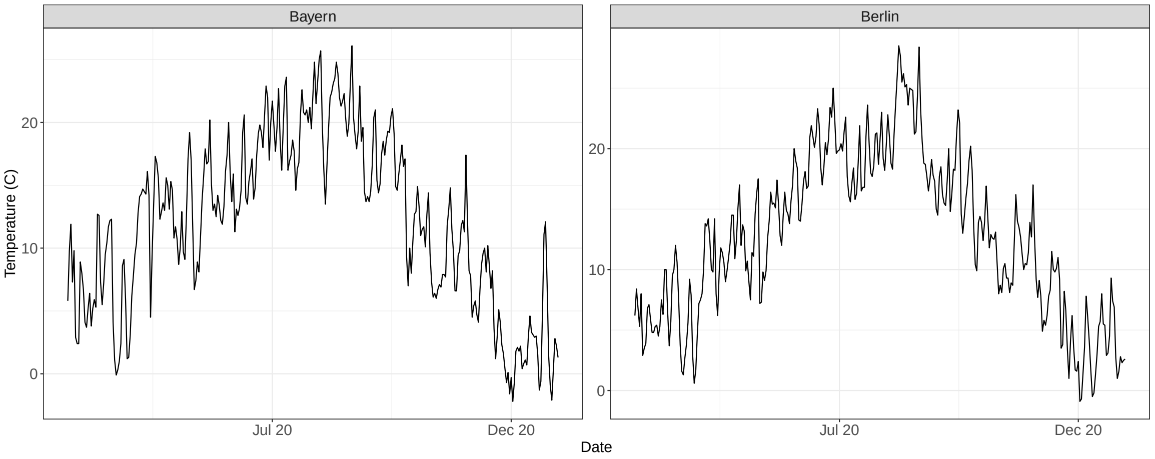


Fig. 12: **Temperature data for Bayern and Berlin, from February 21 , 2020 until December 31, 2020.**

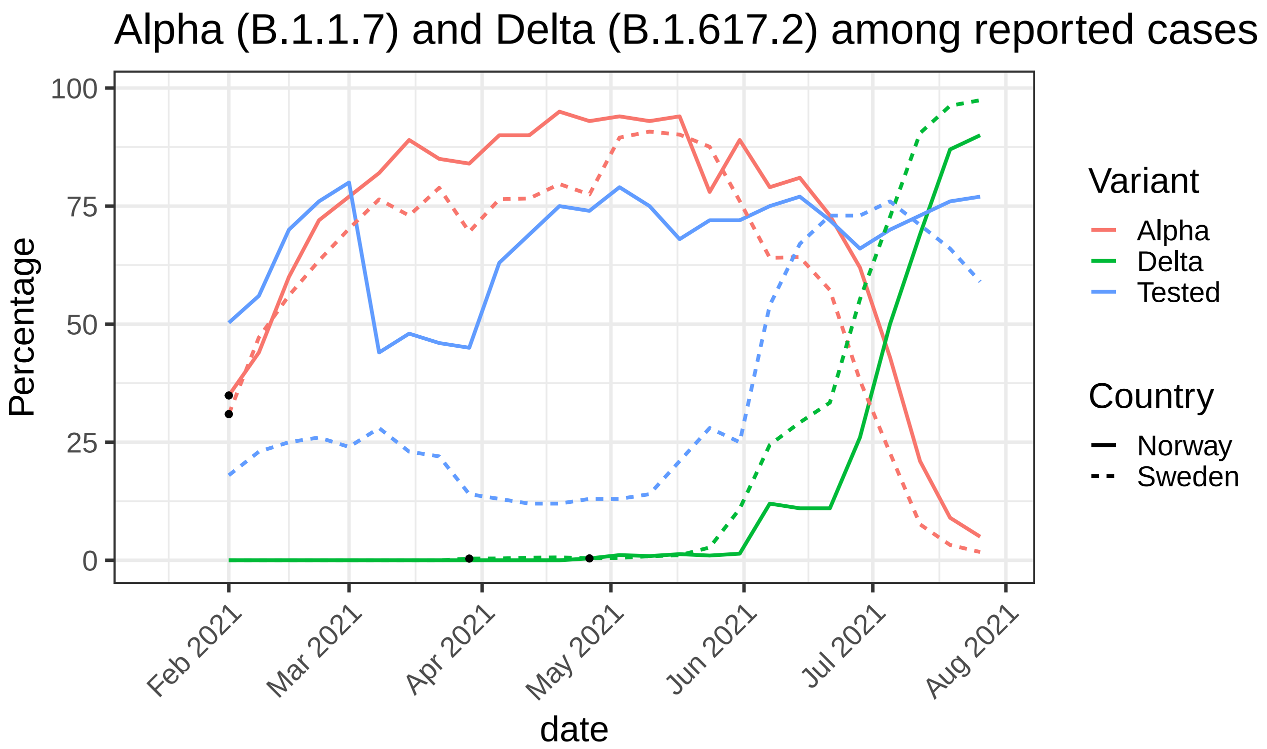


Fig. 13: Proportion of different variants of the sequenced/screened positive tests.Red lines: Alpha, green line: Delta, blue line: proportion of positive tests that are sequenced/screened. Solid line: Norway, dashed line: Sweden.

Table 2: Risk of being hospitalised, given infection, for age groups defined in the vaccination data.

| Country | Age group | Risk (%) |
|---|---|---|
| Norway | 0-15 | 0.1 |
| Norway | 16-44 | 0.5 |
| Norway | 45-54 | 1.9 |
| Norway | 55-64 | 3.7 |
| Norway | 65-74 | 6.7 |
| Norway | 75-84 | 13.1 |
| Norway | 85+ | 24.8 |
| Sweden | 0-11 | 0.1 |
| Sweden | 12-15 | 0.2 |
| Sweden | 16-17 | 0.2 |
| Sweden | 18-29 | 0.37 |
| Sweden | 30-39 | 0.7 |
| Sweden | 40-49 | 1.3 |
| Sweden | 50-59 | 2.5 |
| Sweden | 60-69 | 4.7 |
| Sweden | 70-79 | 9.0 |
| Sweden | 80-89 | 16.9 |
| Sweden | 90+ | 34.1 |

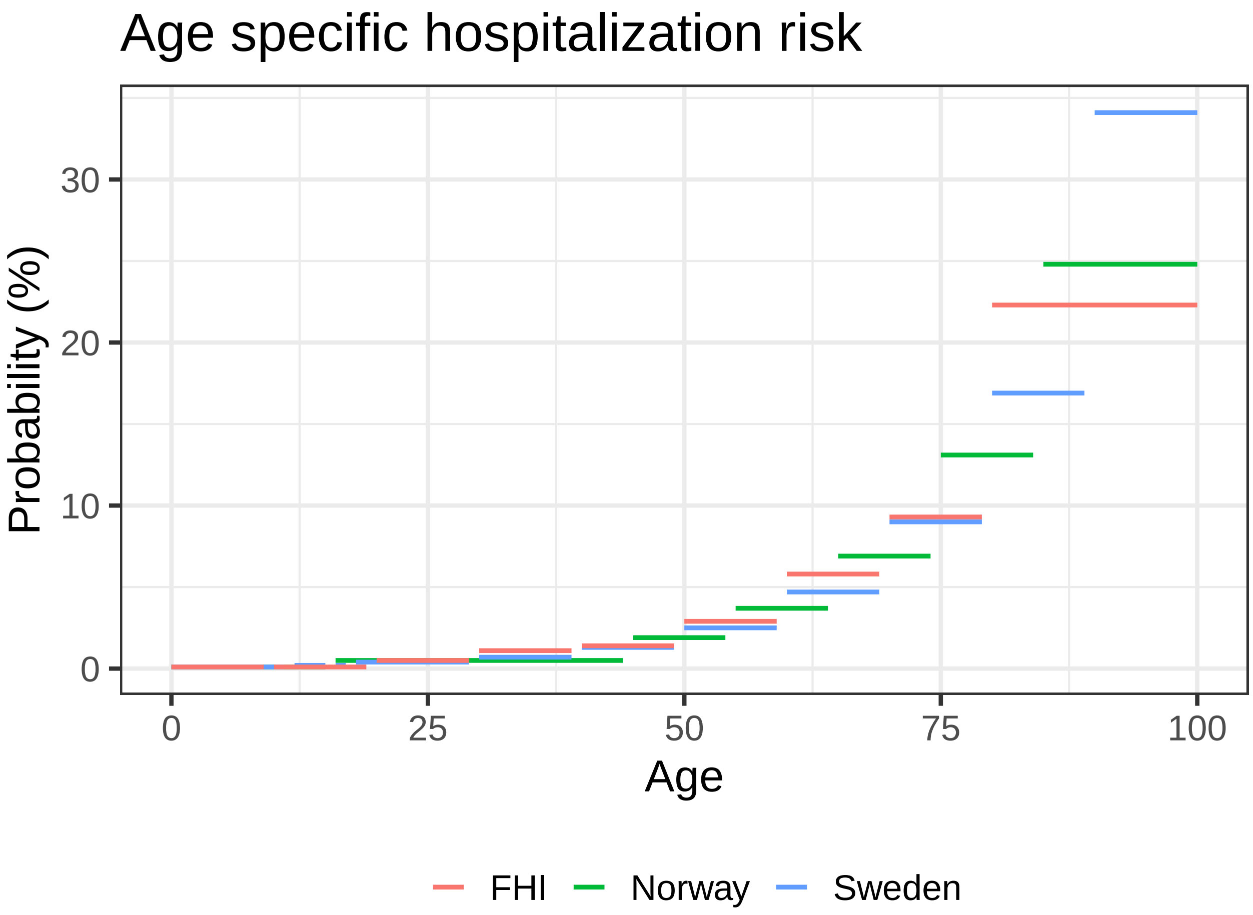


Fig. 14: **Age specific hospitalization risk given infection.** Red lines denote age groups defined by FHI, Norway, green lines denote age groups of the Norwegian vaccination data and blue lines denote age groups of the Swedish vaccination data.

| | County | Share daily inf. contacts (mean) | Share secondary infections |
|---|---|---|---|
| 1 | Agder | 0.430 (0.260-0.588) | 0.296 (0.138-0.466) |
| 2 | Innlandet | 0.432 (0.282-0.561) | 0.295 (0.155-0.430) |
| 3 | Møre og Romsdal | 0.460 (0.276-0.645) | 0.329 (0.145-0.550) |
| 4 | Nordland | 0.537 (0.324-0.755) | 0.423 (0.191-0.706) |
| 5 | Oslo | 0.579 (0.474-0.670) | 0.482 (0.375-0.579) |
| 6 | Rogaland | 0.275 (0.157-0.396) | 0.136 (0.058-0.228) |
| 7 | Troms og Finnmark | 0.381 (0.218-0.544) | 0.240 (0.101-0.405) |
| 8 | Tröndelag | 0.346 (0.193-0.497) | 0.198 (0.081-0.338) |
| 9 | Vestfold og Telemark | 0.373 (0.223-0.515) | 0.229 (0.103-0.370) |
| 10 | Vestland | 0.331 (0.192-0.465) | 0.185 (0.081-0.302) |
| 11 | Viken | 0.533 (0.421-0.625) | 0.428 (0.314-0.527) |
| 12 | Blekinge | 0.584 (0.475-0.679) | 0.479 (0.355-0.585) |
| 13 | Dalarna | 0.625 (0.542-0.694) | 0.568 (0.478-0.643) |
| 14 | Gotland | 0.538 (0.395-0.662) | 0.412 (0.260-0.558) |
| 15 | Gävleborg | 0.593 (0.494-0.676) | 0.518 (0.404-0.613) |
| 16 | Halland | 0.579 (0.470-0.672) | 0.487 (0.366-0.595) |
| 17 | Jämtland | 0.544 (0.430-0.643) | 0.453 (0.324-0.571) |
| 18 | Jönköping | 0.684 (0.605-0.750) | 0.638 (0.544-0.715) |
| 19 | Kalmar | 0.569 (0.467-0.664) | 0.470 (0.352-0.579) |
| 20 | Kronoberg | 0.693 (0.604-0.767) | 0.636 (0.533-0.721) |
| 21 | Norrbotten | 0.548 (0.438-0.642) | 0.455 (0.332-0.558) |
| 22 | Skåne | 0.565 (0.474-0.639) | 0.467 (0.374-0.547) |
| 23 | Stockholm | 0.652 (0.590-0.705) | 0.619 (0.554-0.675) |
| 24 | Södermansland | 0.618 (0.533-0.692) | 0.562 (0.466-0.647) |
| 25 | Uppsala | 0.690 (0.622-0.744) | 0.654 (0.579-0.714) |
| 26 | Värmland | 0.623 (0.533-0.700) | 0.563 (0.462-0.649) |
| 27 | Västerbotten | 0.675 (0.554-0.770) | 0.613 (0.468-0.730) |
| 28 | Västernorrland | 0.653 (0.574-0.719) | 0.588 (0.499-0.664) |
| 29 | Västmanland | 0.599 (0.507-0.681) | 0.531 (0.428-0.627) |
| 30 | Västra Götaland | 0.733 (0.681-0.778) | 0.702 (0.645-0.750) |
| 31 | Örebro | 0.545 (0.446-0.635) | 0.450 (0.343-0.550) |
| 32 | Östergötland | 0.646 (0.578-0.706) | 0.602 (0.527-0.667) |

Table 3: **SARS-CoV-2 transmission dynamics explained by COVID-19 community mobility reports.** This table shows: (i) the average fraction of (potentially infectious) daily contacts explained by activity levels within settings captured by the COVID-19 community mobility report. These contacts lead to a secondary infection if an infected meets a susceptible. (ii) The model based share of secondary infections (over the whole observation period) assigned to settings captured by the four mobility reports considered in the model. Shown are the posterior median and 90%-credible intervals per region.

| | County | Share daily inf. contacts (mean) | Share secondary infections |
|---|---|---|---|
| 1 | Berlin | 0.693 (0.615,0.759) | 0.726 (0.638,0.795) |
| 2 | Bayern | 0.642 (0.575,0.700) | 0.618 (0.543,0.679) |

Table 4: **SARS-CoV-2 transmission dynamics explained by COVID-19 community mobility reports.** This table shows: (i) the average fraction of (potentially infectious) daily contacts explained by activity levels within settings captured by the COVID-19 community mobility report. These contacts lead to a secondary infection if an infected meets a susceptible. (ii) The model based share of secondary infections (over the whole observation period) assigned to settings captured by the four mobility reports considered in the model. Shown are the posterior median and 90%-credible intervals per region, for the German regions Bayern and Berlin.

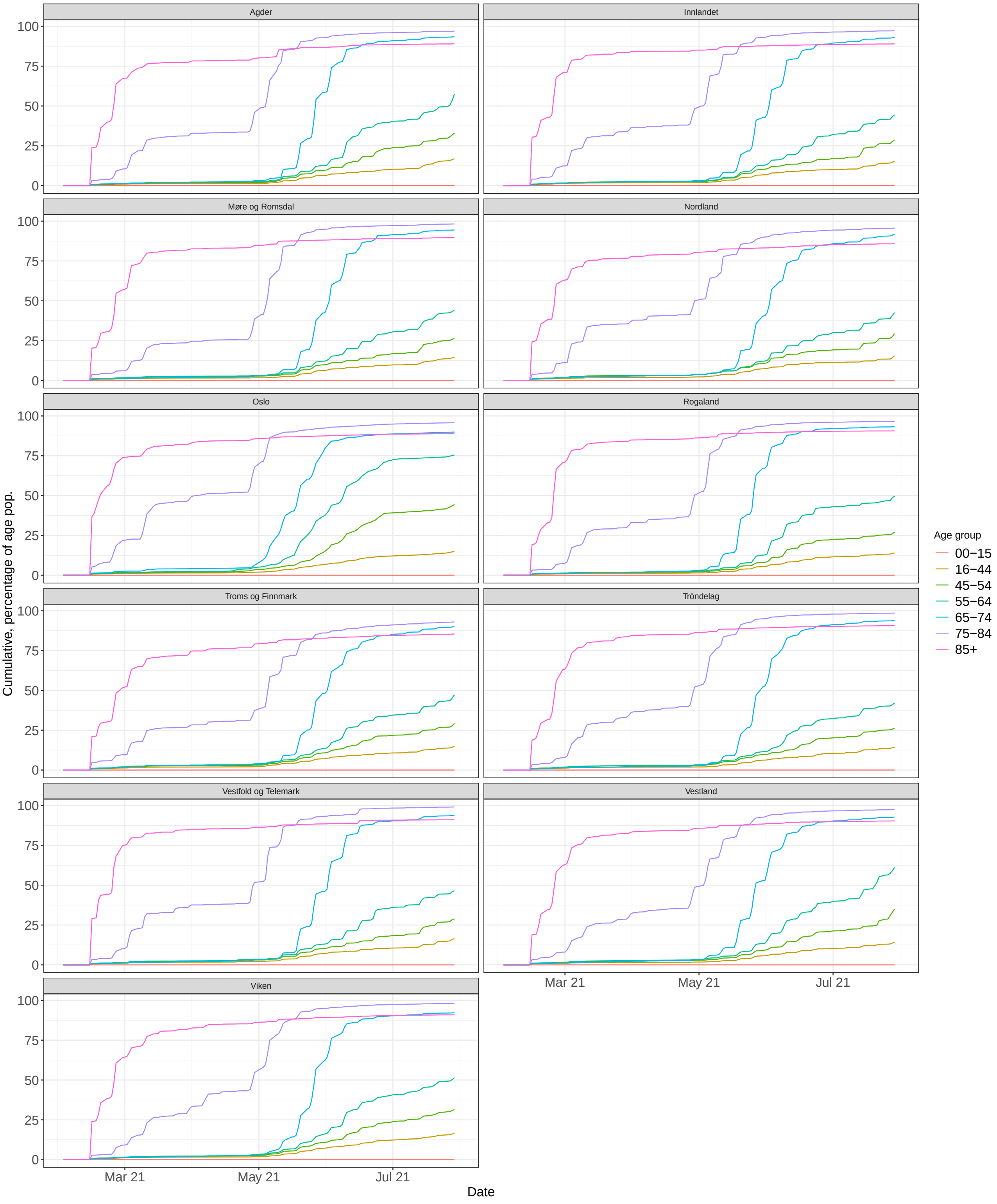


Fig. 15: **Vaccination by age group and region, Norway.** Cumulative number of individuals with 2 vaccine doses, as percentage of age stratified population, for all Norwegian regions.

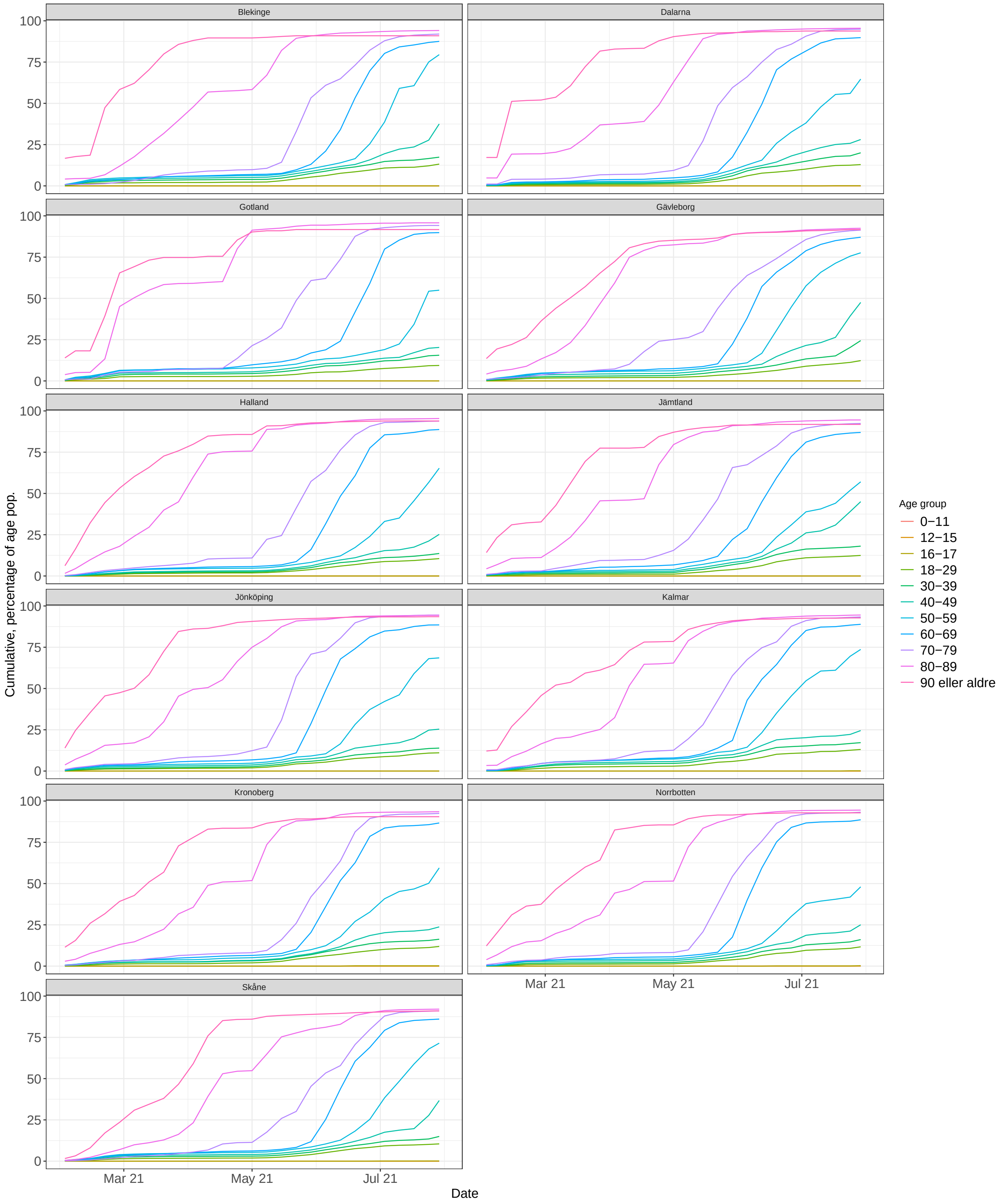


Fig. 16: **Vaccination by age group and region, Sweden.** Cumulative number of individuals with 2 vaccine doses, as percentage of age stratified population, for 11 Swedish regions.

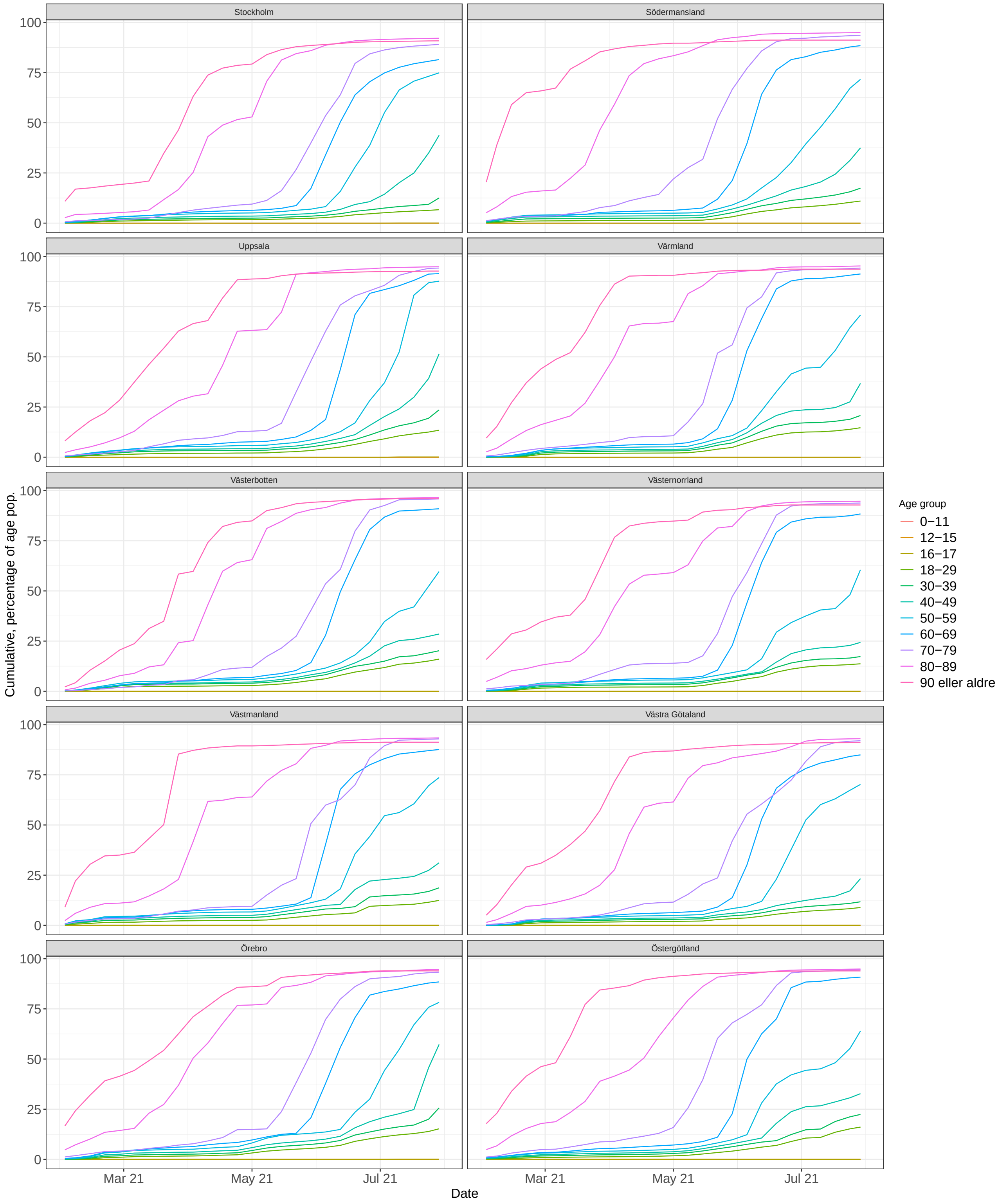


Fig. 17: **Vaccination by age group and region, Sweden.** Cumulative number of individuals with 2 vaccine doses, as percentage of age stratified population, for 10 Swedish regions.

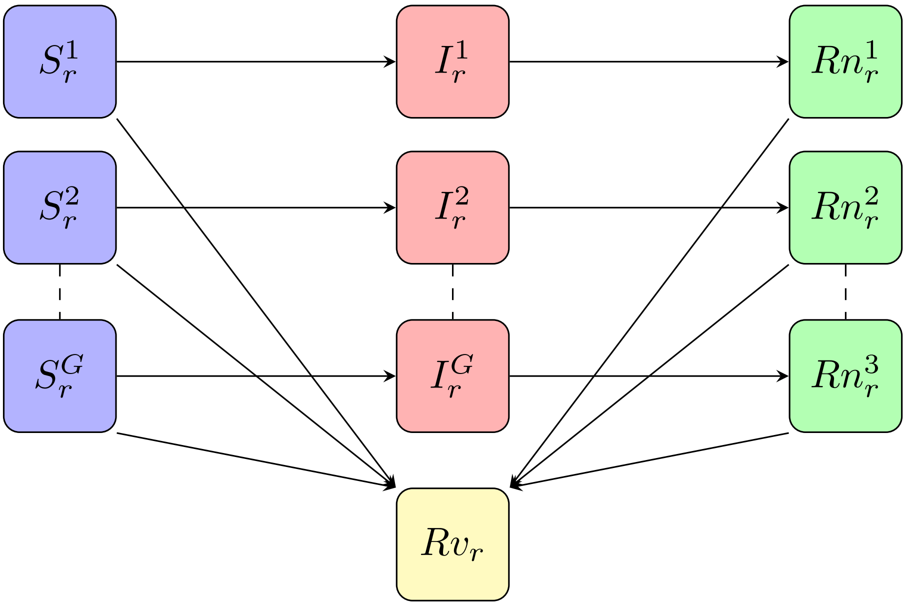


Fig. 18: **State space model for disease spread.** Schematic visualization of the metapopulation model used to model disease spread in a region.

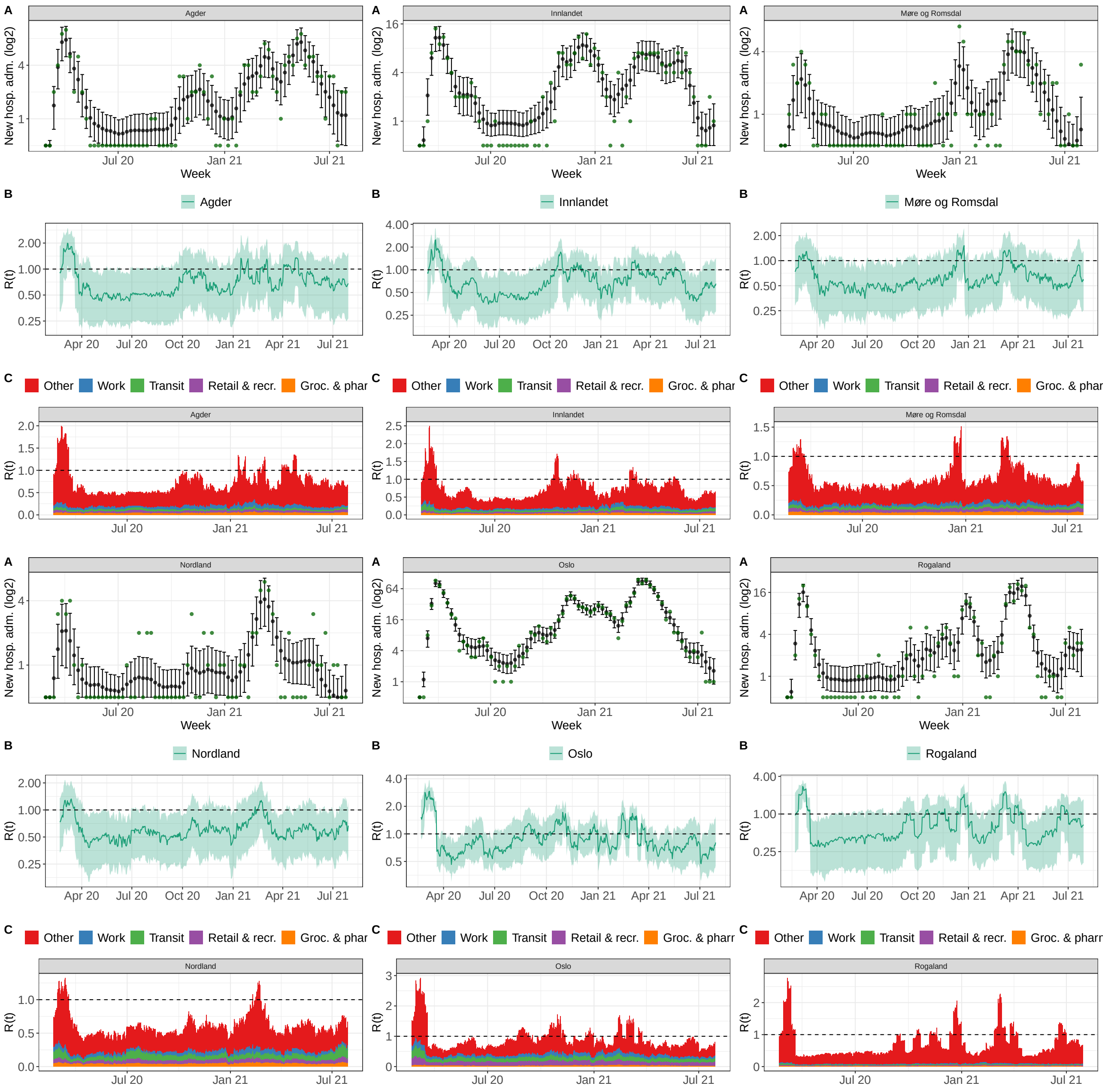


Fig. 19: Model fit Norwegian regions A-R

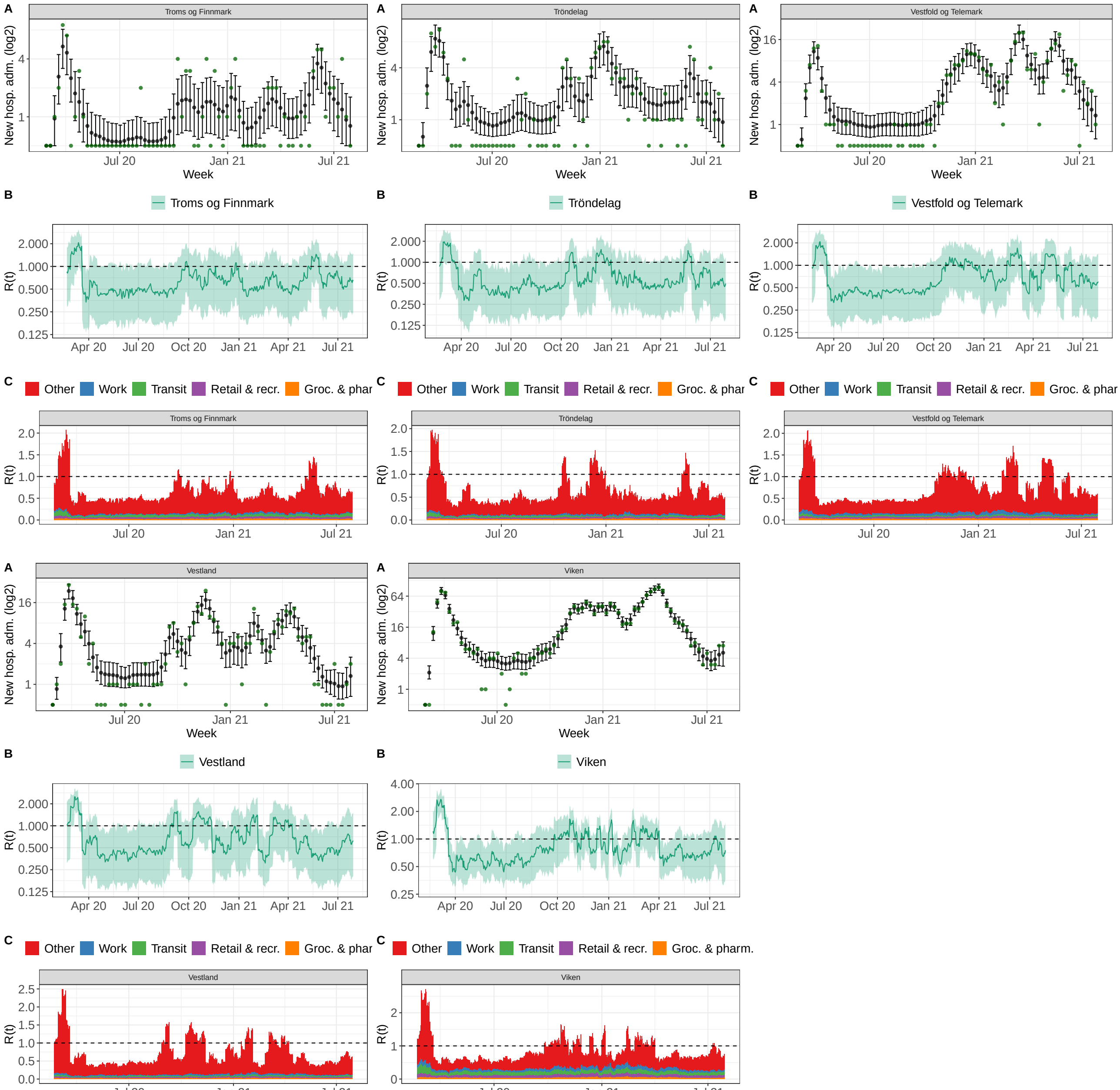


Fig. 20: Model fit Norwegian regions S-Z

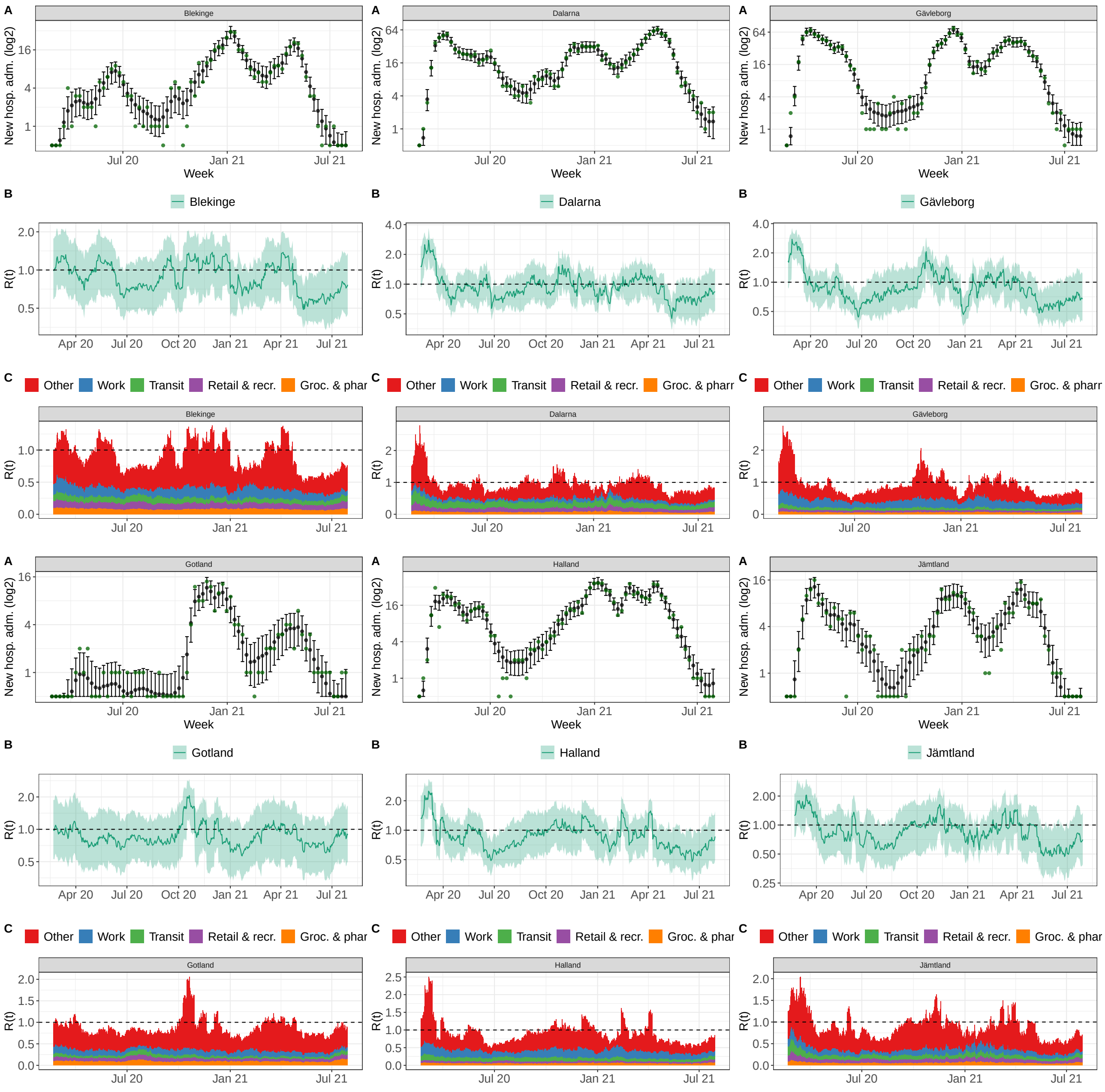


Fig. 21: Model fit Swedish regions A-J

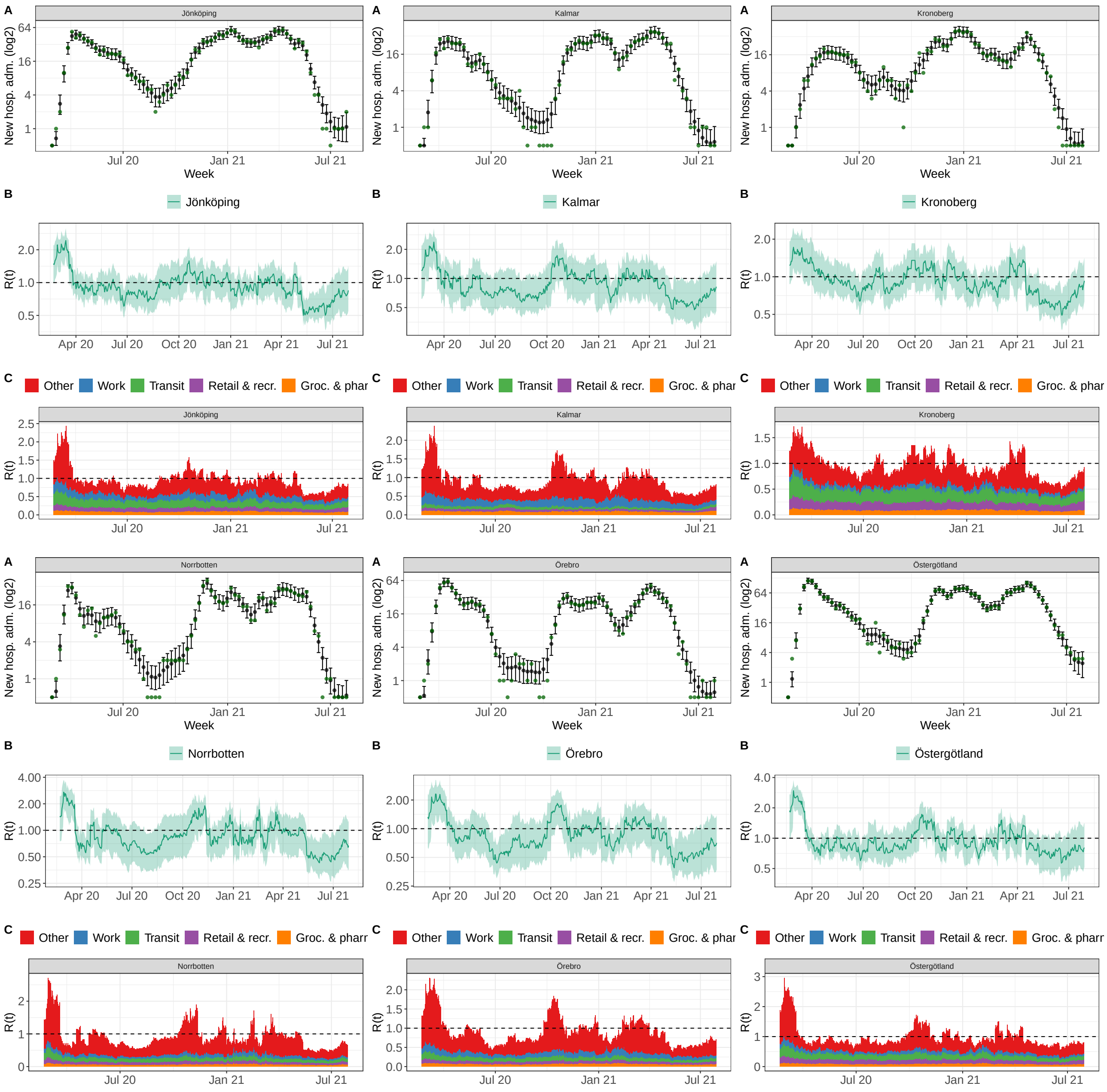


Fig. 22: Model fit Swedish regions K-O

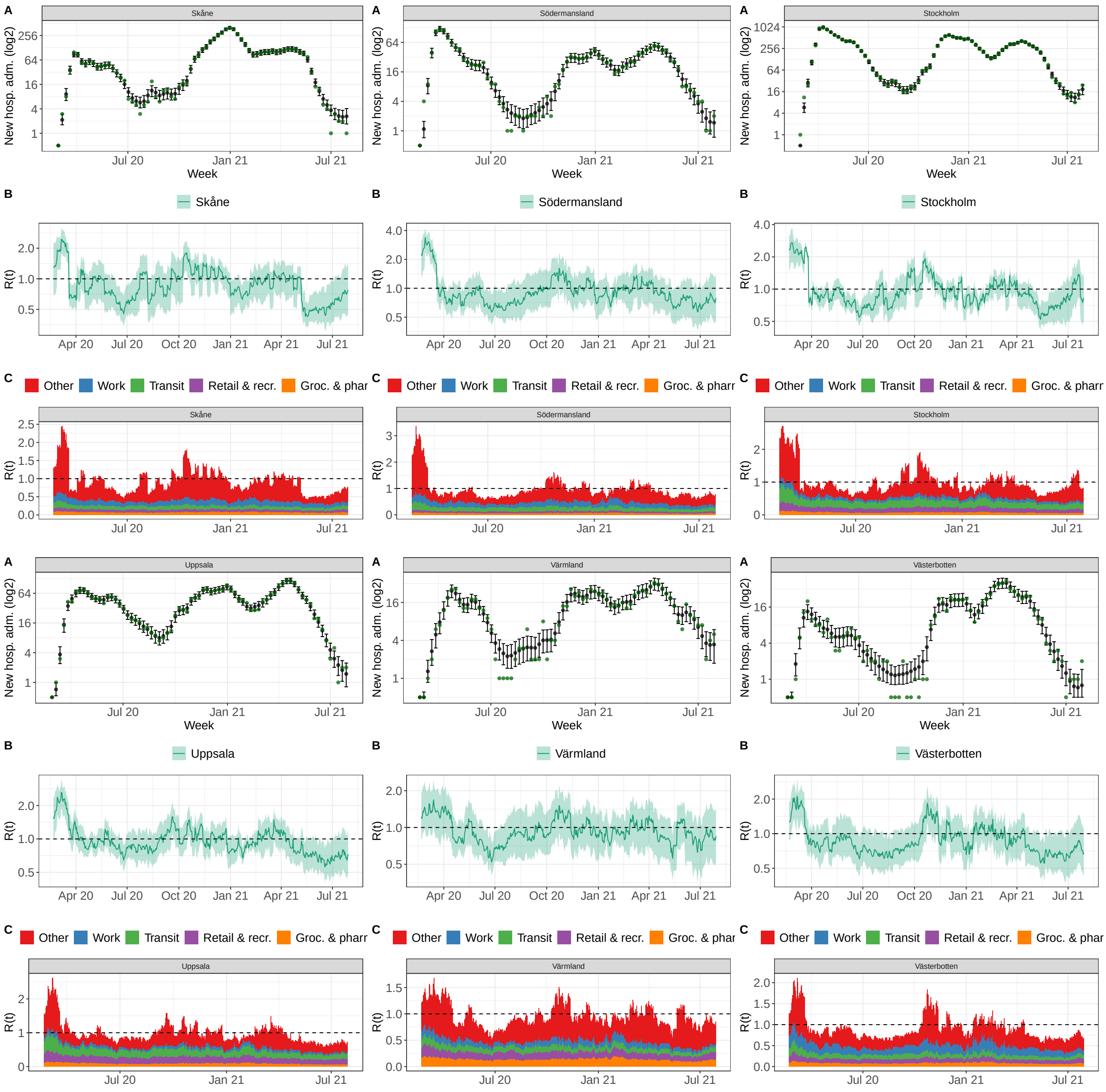


Fig. 23: Model fit Swedish regions P-V

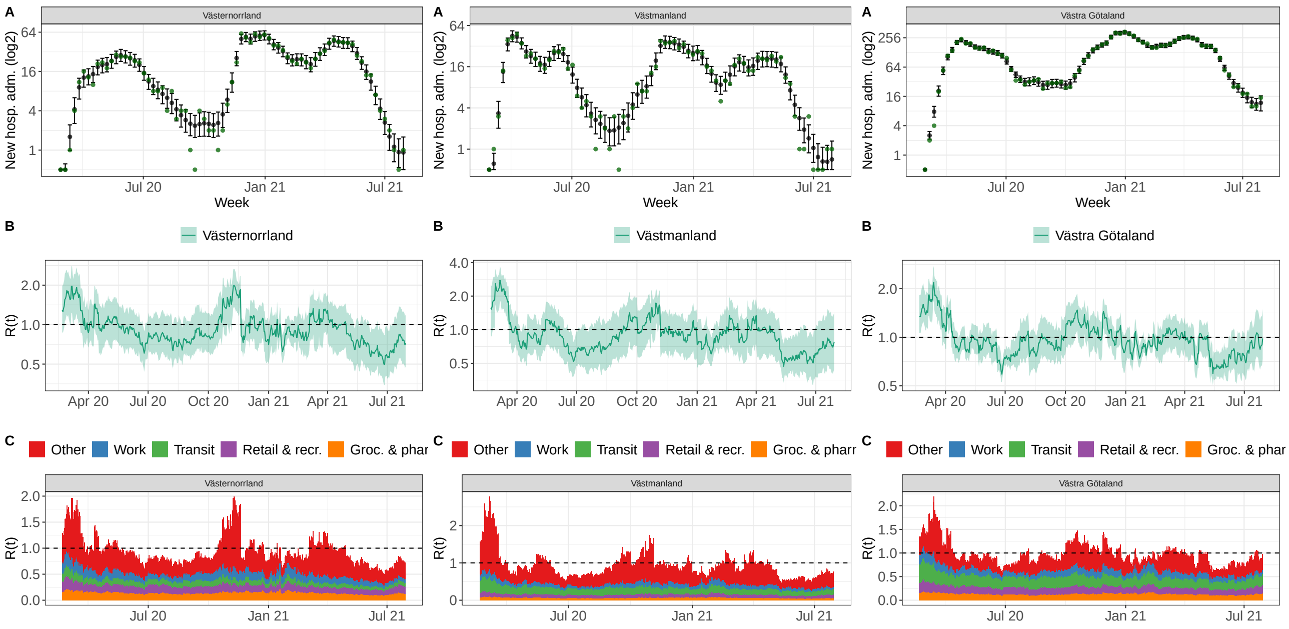


Fig. 24: Model fit Swedish regions V (cont.) - Z

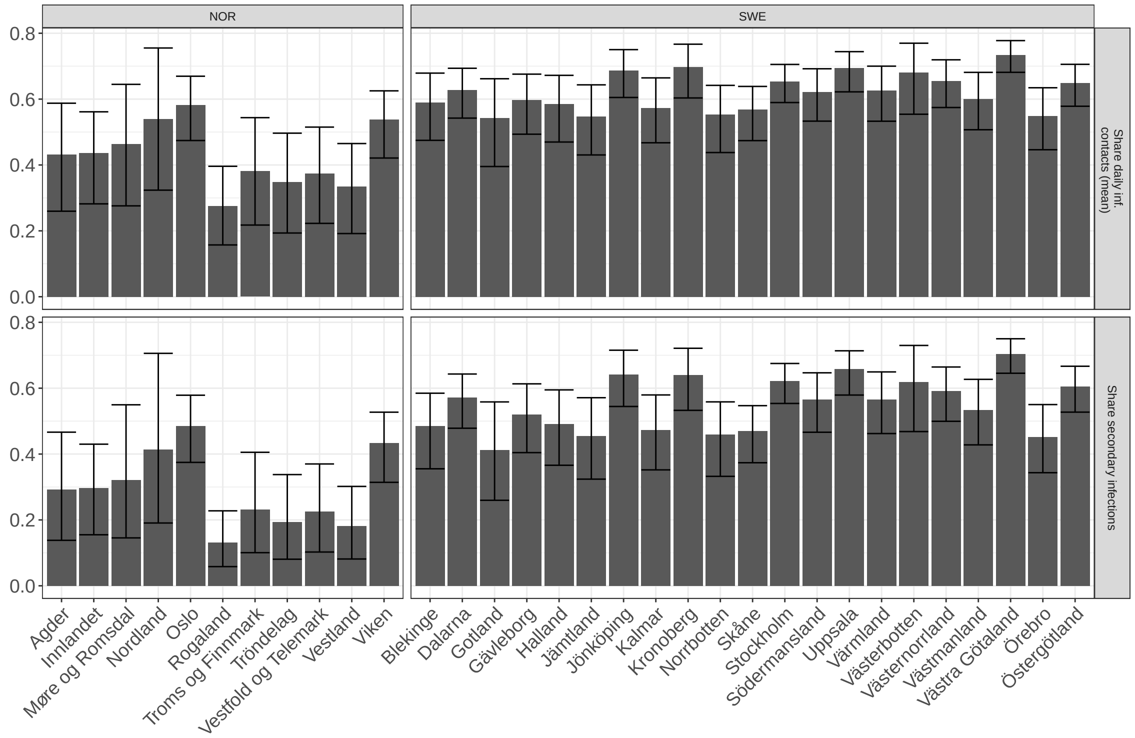


Fig. 25: **SARS-CoV-2 transmission dynamics explained by COVID-19 community mobility reports:** (i) the average fraction of (potentially infectious) daily contacts explained by activity levels within settings captured by the COVID-19 community mobility report. These contacts lead to a secondary infection if an infected meets a susceptible. (ii) The model based share of secondary infections (over the whole observation period) assigned to settings captured by the four mobility reports considered in the model. Shown are the posterior median and 90%-credible intervals per region.

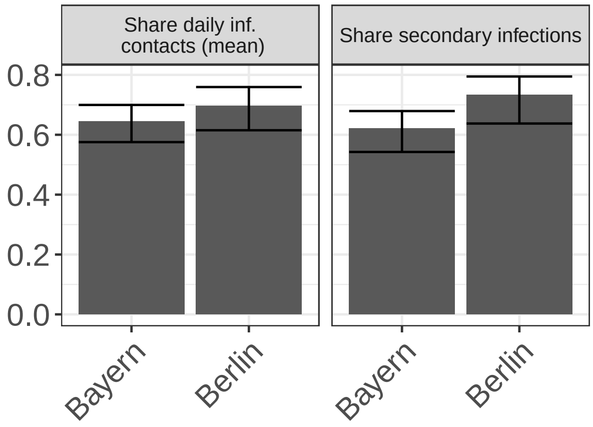


Fig. 26: **SARS-CoV-2 transmission dynamics explained by COVID-19 community mobility reports:** (i) the average fraction of (potentially infectious) daily contacts explained by activity levels within settings captured by the COVID-19 community mobility report. These contacts lead to a secondary infection if an infected meets a susceptible. (ii) The model based share of secondary infections (over the whole observation period) assigned to settings captured by the four mobility reports considered in the model. Shown are the posterior median and 90%-credible intervals per region.

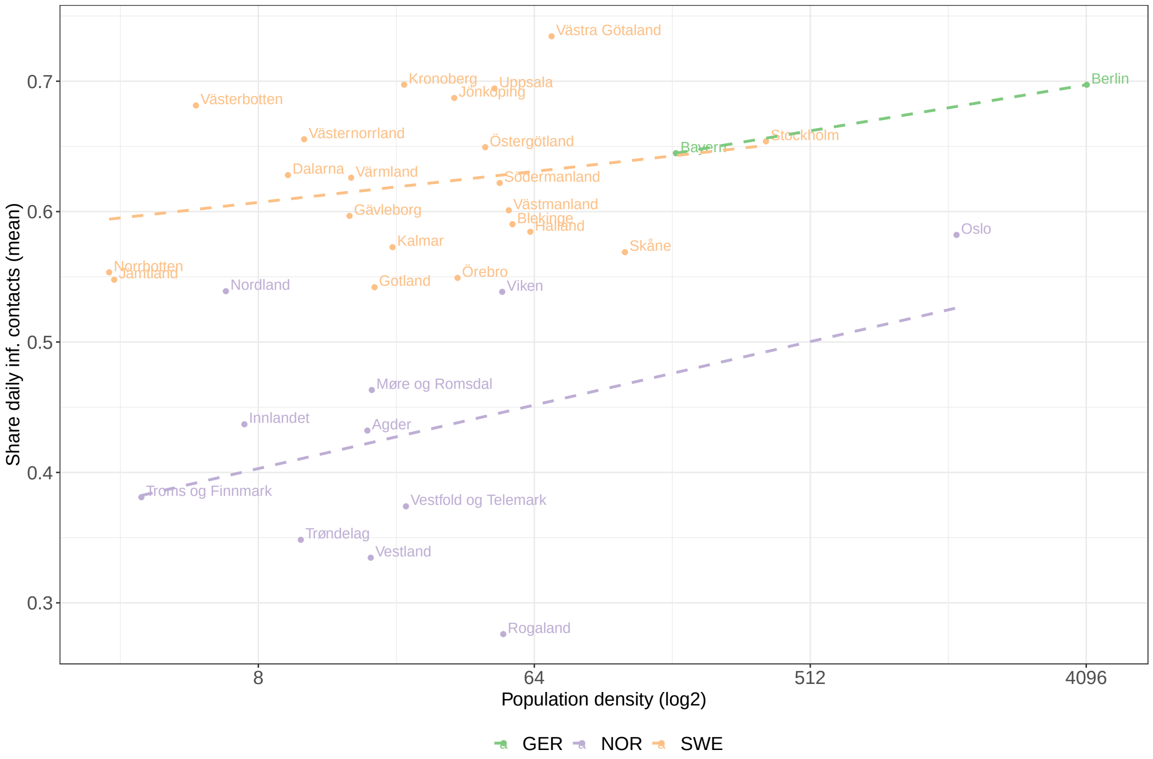


Fig. 27: **Association of population density with share of SARS-CoV-2 transmission dynamics explained by COVID-19 community mobility reports:** Scatterplot of the population density (number of inhabitants per $km^2$) and the average fraction of (potentially infectious) daily contacts explained by activity levels within settings captured by the COVID-19 community mobility reports per region and their linear association per country (dotted line). Note that population density is on a log-scale and that the modelling period for the two German regions are shorter than for the Norwegian and Swedish regions.

| County | name | Groc. & pharm | Retail & recr. | Transit | Work |
|---|---|---|---|---|---|
| Agder | Coef. | 0.0082 (0.00069-0.029) | 0.0086 (0.00063-0.031) | 0.0087 (0.00082-0.030) | 0.0154 (0.00122-0.050) |
| | Share | 0.17 (0.014-0.56) | 0.18 (0.014-0.58) | 0.18 (0.017-0.58) | 0.33 (0.033-0.73) |
| Innlandet | Coef. | 0.0077 (0.00065-0.027) | 0.0084 (0.00066-0.030) | 0.0186 (0.00151-0.058) | 0.0127 (0.00098-0.041) |
| | Share | 0.13 (0.011-0.50) | 0.15 (0.012-0.55) | 0.34 (0.035-0.75) | 0.23 (0.018-0.64) |
| Møre og Romsdal | Coef. | 0.0082 (0.00075-0.031) | 0.0097 (0.00068-0.035) | 0.0076 (0.00068-0.027) | 0.0113 (0.00086-0.042) |
| | Share | 0.19 (0.017-0.59) | 0.22 (0.017-0.63) | 0.17 (0.016-0.56) | 0.26 (0.023-0.70) |
| Nordland | Coef. | 0.0080 (0.00061-0.031) | 0.0091 (0.00085-0.033) | 0.0161 (0.00130-0.050) | 0.0119 (0.00093-0.045) |
| | Share | 0.15 (0.011-0.53) | 0.17 (0.016-0.57) | 0.30 (0.030-0.71) | 0.23 (0.020-0.63) |
| Oslo | Coef. | 0.0087 (0.00061-0.035) | 0.0399 (0.00352-0.101) | 0.0500 (0.00461-0.127) | 0.0341 (0.00342-0.092) |
| | Share | 0.058 (0.0038-0.26) | 0.273 (0.0230-0.67) | 0.338 (0.0338-0.75) | 0.232 (0.0223-0.61) |
| Rogaland | Coef. | 0.0038 (0.00032-0.015) | 0.0047 (0.00033-0.018) | 0.0064 (0.00051-0.023) | 0.0050 (0.00047-0.019) |
| | Share | 0.16 (0.013-0.55) | 0.19 (0.016-0.62) | 0.27 (0.025-0.69) | 0.21 (0.021-0.64) |
| Troms og Finnmark | Coef. | 0.0053 (0.00047-0.020) | 0.0084 (0.00063-0.032) | 0.0122 (0.00105-0.045) | 0.0067 (0.00051-0.027) |
| | Share | 0.13 (0.011-0.49) | 0.22 (0.018-0.63) | 0.32 (0.031-0.73) | 0.17 (0.013-0.59) |
| Tröndelag | Coef. | 0.0048 (0.00044-0.019) | 0.0061 (0.00046-0.024) | 0.0110 (0.00079-0.041) | 0.0067 (0.00045-0.026) |
| | Share | 0.14 (0.013-0.53) | 0.17 (0.014-0.56) | 0.33 (0.029-0.75) | 0.20 (0.013-0.63) |
| Vestfold og Telemark | Coef. | 0.0059 (0.00048-0.022) | 0.0077 (0.00068-0.028) | 0.0071 (0.00057-0.028) | 0.0132 (0.00112-0.042) |
| | Share | 0.15 (0.012-0.53) | 0.19 (0.017-0.61) | 0.18 (0.014-0.57) | 0.33 (0.035-0.76) |
| Vestland | Coef. | 0.0049 (0.00041-0.018) | 0.0065 (0.00055-0.025) | 0.0061 (0.00056-0.022) | 0.0085 (0.00056-0.031) |
| | Share | 0.16 (0.013-0.53) | 0.21 (0.017-0.65) | 0.19 (0.018-0.59) | 0.28 (0.020-0.70) |
| Viken | Coef. | 0.0089 (0.00072-0.034) | 0.0166 (0.00123-0.050) | 0.0369 (0.00364-0.091) | 0.0216 (0.00198-0.063) |
| | Share | 0.089 (0.0066-0.40) | 0.167 (0.0115-0.55) | 0.391 (0.0450-0.77) | 0.225 (0.0202-0.63) |
| Blekinge | Coef. | 0.017 (0.0013-0.056) | 0.018 (0.0014-0.058) | 0.019 (0.0015-0.057) | 0.037 (0.0038-0.086) |
| | Share | 0.16 (0.012-0.55) | 0.17 (0.013-0.58) | 0.18 (0.015-0.52) | 0.35 (0.039-0.73) |
| Dalarna | Coef. | 0.016 (0.0010-0.054) | 0.029 (0.0029-0.080) | 0.047 (0.0051-0.106) | 0.027 (0.0028-0.069) |
| | Share | 0.11 (0.008-0.42) | 0.22 (0.022-0.60) | 0.36 (0.042-0.74) | 0.20 (0.022-0.52) |
| Gotland | Coef. | 0.018 (0.00166-0.052) | 0.013 (0.00096-0.044) | 0.013 (0.00116-0.042) | 0.028 (0.00262-0.073) |
| | Share | 0.22 (0.020-0.61) | 0.16 (0.011-0.55) | 0.16 (0.014-0.49) | 0.34 (0.039-0.71) |
| Gävleborg | Coef. | 0.013 (0.00098-0.042) | 0.013 (0.00123-0.047) | 0.020 (0.00189-0.065) | 0.060 (0.01083-0.108) |
| | Share | 0.10 (0.0083-0.40) | 0.11 (0.0105-0.43) | 0.17 (0.0157-0.55) | 0.51 (0.0991-0.82) |
| Halland | Coef. | 0.012 (0.00098-0.042) | 0.010 (0.00069-0.036) | 0.026 (0.00274-0.073) | 0.051 (0.00811-0.096) |
| | Share | 0.109 (0.0086-0.42) | 0.093 (0.0063-0.35) | 0.241 (0.0271-0.63) | 0.475 (0.0895-0.80) |
| Jämtland | Coef. | 0.013 (0.00092-0.046) | 0.020 (0.00171-0.061) | 0.029 (0.00244-0.082) | 0.032 (0.00359-0.076) |
| | Share | 0.12 (0.0088-0.45) | 0.19 (0.0151-0.56) | 0.27 (0.0257-0.66) | 0.29 (0.0343-0.68) |
| Jönköping | Coef. | 0.019 (0.0016-0.059) | 0.024 (0.0021-0.073) | 0.056 (0.0084-0.118) | 0.043 (0.0051-0.097) |
| | Share | 0.12 (0.010-0.41) | 0.15 (0.014-0.50) | 0.36 (0.057-0.70) | 0.28 (0.035-0.60) |
| Kalmar | Coef. | 0.017 (0.0017-0.053) | 0.011 (0.0011-0.037) | 0.017 (0.0012-0.054) | 0.047 (0.0065-0.090) |
| | Share | 0.17 (0.016-0.56) | 0.11 (0.010-0.39) | 0.16 (0.013-0.52) | 0.46 (0.078-0.78) |
| Kronoberg | Coef. | 0.020 (0.0013-0.065) | 0.035 (0.0029-0.092) | 0.065 (0.0098-0.138) | 0.022 (0.0025-0.067) |
| | Share | 0.12 (0.0076-0.45) | 0.22 (0.0179-0.61) | 0.42 (0.0688-0.77) | 0.14 (0.0160-0.43) |
| Norrbotten | Coef. | 0.013 (0.0010-0.050) | 0.019 (0.0016-0.059) | 0.034 (0.0032-0.091) | 0.023 (0.0023-0.064) |
| | Share | 0.13 (0.0093-0.51) | 0.18 (0.0149-0.58) | 0.33 (0.0347-0.74) | 0.22 (0.0224-0.60) |
| Skåne | Coef. | 0.015 (0.0012-0.050) | 0.016 (0.0017-0.050) | 0.030 (0.0028-0.081) | 0.036 (0.0041-0.082) |
| | Share | 0.13 (0.010-0.49) | 0.14 (0.014-0.46) | 0.27 (0.028-0.66) | 0.33 (0.040-0.71) |
| Stockholm | Coef. | 0.018 (0.0012-0.060) | 0.040 (0.0040-0.101) | 0.073 (0.0084-0.153) | 0.031 (0.0023-0.094) |
| | Share | 0.098 (0.0065-0.37) | 0.220 (0.0214-0.59) | 0.409 (0.0515-0.77) | 0.175 (0.0129-0.53) |
| Södermansland | Coef. | 0.014 (0.0012-0.050) | 0.014 (0.0011-0.050) | 0.045 (0.0049-0.106) | 0.048 (0.0057-0.099) |
| | Share | 0.10 (0.0089-0.41) | 0.10 (0.0079-0.42) | 0.33 (0.0406-0.71) | 0.35 (0.0479-0.72) |
| Uppsala | Coef. | 0.021 (0.0015-0.065) | 0.047 (0.0062-0.109) | 0.072 (0.0082-0.154) | 0.024 (0.0026-0.071) |
| | Share | 0.11 (0.0083-0.39) | 0.26 (0.0327-0.64) | 0.40 (0.0515-0.76) | 0.13 (0.0138-0.40) |
| Värmland | Coef. | 0.029 (0.0024-0.077) | 0.031 (0.0030-0.081) | 0.026 (0.0023-0.079) | 0.026 (0.0023-0.074) |
| | Share | 0.23 (0.019-0.63) | 0.24 (0.024-0.61) | 0.21 (0.018-0.59) | 0.20 (0.019-0.55) |
| Västerbotten | Coef. | 0.017 (0.0015-0.056) | 0.024 (0.0020-0.070) | 0.031 (0.0032-0.088) | 0.051 (0.0083-0.108) |
| | Share | 0.12 (0.010-0.45) | 0.17 (0.015-0.53) | 0.23 (0.025-0.59) | 0.36 (0.064-0.72) |
| Västernorrland | Coef. | 0.028 (0.0022-0.075) | 0.031 (0.0043-0.079) | 0.026 (0.0024-0.076) | 0.035 (0.0035-0.084) |
| | Share | 0.20 (0.015-0.58) | 0.23 (0.032-0.57) | 0.19 (0.017-0.54) | 0.26 (0.027-0.60) |
| Västmanland | Coef. | 0.013 (0.0009-0.047) | 0.019 (0.0017-0.058) | 0.055 (0.0064-0.123) | 0.026 (0.0025-0.070) |
| | Share | 0.10 (0.0065-0.42) | 0.15 (0.0132-0.51) | 0.44 (0.0638-0.79) | 0.20 (0.0203-0.56) |
| Västra Götaland | Coef. | 0.027 (0.0023-0.076) | 0.035 (0.0041-0.092) | 0.066 (0.0112-0.132) | 0.035 (0.0046-0.080) |
| | Share | 0.15 (0.013-0.46) | 0.20 (0.022-0.54) | 0.37 (0.069-0.69) | 0.20 (0.028-0.45) |
| Örebro | Coef. | 0.014 (0.0011-0.048) | 0.016 (0.0012-0.053) | 0.029 (0.0031-0.082) | 0.032 (0.0036-0.076) |
| | Share | 0.13 (0.010-0.49) | 0.15 (0.011-0.52) | 0.28 (0.031-0.69) | 0.30 (0.035-0.69) |
| Östergötland | Coef. | 0.019 (0.0016-0.060) | 0.031 (0.0029-0.083) | 0.056 (0.0051-0.128) | 0.030 (0.0028-0.083) |
| | Share | 0.12 (0.010-0.43) | 0.20 (0.018-0.59) | 0.37 (0.037-0.74) | 0.20 (0.020-0.55) |

Table 5: Estimated coefficients $\hat{\phi}_{k,r}$ as well as their relative size per region and associated 90% credible intervals for Norwegian and Swedish regions.

| County | name | Groc. & pharm | Retail & recr. | Transit | Work |
|---|---|---|---|---|---|
| Bayern | Coef. | 0.015 (0.0011-0.048) | 0.065 (0.0099-0.130) | 0.056 (0.0064-0.131) | 0.019 (0.0016-0.064) |
| | Share | 0.089 (0.0063-0.31) | 0.381 (0.0581-0.76) | 0.329 (0.0405-0.72) | 0.109 (0.0089-0.39) |
| Berlin | Coef. | 0.013 (0.0010-0.048) | 0.125 (0.0351-0.202) | 0.046 (0.0047-0.129) | 0.026 (0.0021-0.084) |
| | Share | 0.056 (0.0044-0.23) | 0.557 (0.1726-0.83) | 0.207 (0.0206-0.58) | 0.112 (0.0095-0.39) |

Table 6: Estimated coefficients $\hat{\phi}_{k,r}$ as well as their relative size per region and associated 90% credible intervals for German regions.
